# Temporal dynamics of radiotherapy and chemotherapy response in lower-grade gliomas using causal machine learning

**DOI:** 10.64898/2026.02.28.26347288

**Authors:** Everest Yang, Sparsh Agarwal, Connor J. Kinslow, Simon K. Cheng, Lillian Yang, Eric Wang, Tony J. Wang, Lisa A. Kachnic, David J. Brenner, Igor Shuryak

**Affiliations:** Center for Radiological Research, Columbia University Irving Medical Center, New York, NY, USA; Department of Computer Science, Brown University, Providence, RI, USA; Department of Biology, Brown University, Providence, RI, USA; Department of Radiation Oncology, Memorial Sloan Kettering Cancer Center, New York, NY, USA; Department of Radiation Oncology, Columbia University Irving Medical Center, New York, NY, USA

**Author notes:** Corresponding Author: Everest Yang.

## Abstract

Lower-grade gliomas (World Health Organization [WHO] grades 2-3) exhibit variable treatment responses, yet clinical decisions remain guided by population-level trial results. Standard causal survival forests estimate treatment effects at individual time horizons but lack methodology to synthesize these into interpretable temporal trajectories. Here, we apply the Causal Analysis of Survival Trajectories (CAST) framework, a recently developed extension of causal survival forests that synthesizes horizon-specific causal effect estimates into smooth temporal curves while accounting for between-horizon covariances via bootstrap estimation and Ledoit-Wolf shrinkage. We apply CAST to estimate time-varying, heterogeneous effects of radiotherapy and chemotherapy in 776 patients with lower-grade gliomas from The Cancer Genome Atlas (TCGA; n=512) and the Chinese Glioma Genome Atlas (CGGA; n=264), analyzing six treatment-outcome scenarios and adjusting for age, sex, WHO grade, isocitrate dehydrogenase (IDH) mutation status, 1p/19q codeletion, and extent of resection using elastic net propensity scores with overlap weighting. CAST curves reveal that chemotherapy provides consistent, sustained benefits across both cohorts; survival probability gains peak at 0.31 at 72-84 months for TCGA overall survival and 0.46 at 48 months for progression-free survival, with restricted mean survival time gains of 18.4 and 32.5 months at 10 years, respectively. CGGA chemotherapy shows delayed but large positive effects (survival probability peak 0.48 at 108 months).

Radiotherapy effects are mixed, with modest E-values indicating sensitivity to residual confounding by indication. Subgroup CAST curves identify age at diagnosis as the dominant driver of treatment effect heterogeneity (46-56% of splits). All findings are robust to placebo permutation, simulated unobserved confounder, and negative control refutation tests. The CAST framework provides a general-purpose tool for temporal treatment effect visualization applicable beyond neuro-oncology.

## Introduction

Lower-grade gliomas (WHO grades 2-3) are slow-growing but biologically diverse tumors^1,2^. The recent advent of molecular classification has reshaped how they are defined and treated^3,4^. Isocitrate dehydrogenase (IDH) mutations, present in approximately 80% of lower-grade gliomas, are associated with better outcomes, and 1p/19q codeletion in oligodendrogliomas confers especially favorable survival^5,6^. The 2021 World Health Organization (WHO) framework prioritizes IDH status and recognizes that certain IDH-wildtype lower-grade gliomas with molecular features such as Telomerase Reverse Transcriptase (TERT) promoter mutation, Epidermal Growth Factor Receptor (EGFR) amplification, or chromosome 7/10 imbalance may behave as the more aggressive glioblastomas despite lower histological grade^4,7^. Standard therapy for lower-grade gliomas begins with maximal safe resection, followed by adjuvant radiotherapy and/or chemotherapy depending on residual disease and risk factors^8,9^.

Landmark randomized controlled trials have established the efficacy of combined modality treatment in this population. RTOG 9802 demonstrated that adjuvant procarbazine, lomustine, and vincristine (PCV) after radiotherapy improves overall survival in high-risk lower-grade gliomas^10^. RTOG 9402 and EORTC 26951 showed long-term benefits of PCV-based chemoradiation in anaplastic oligodendrogliomas^11,12^. The CATNON trial established the benefit of adjuvant temozolomide in 1p/19q non-codeleted anaplastic gliomas^13^. Although some retrospective data suggest that PCV combined with radiotherapy may be superior to temozolomide-based regimens^14,15^, no head-to-head comparisons have been published; the ongoing CODEL trial^16^ and recently presented ECOG-ACRIN E3F05 data^17^ continue to refine comparative effectiveness. These studies confirm that radiotherapy and chemotherapy confer survival benefits, but their temporal dynamics - when effects emerge, peak, and potentially attenuate - remain poorly understood.

The challenge is compounded by the observational nature of most real-world glioma data, where treatment assignment depends on patient characteristics, institutional practices, and evolving standards of care. While randomized controlled trials remain the gold standard, they are limited by narrow inclusion criteria, ethical constraints on withholding effective treatments, and high costs that restrict long-term follow-up^18^. Observational datasets offer broader representation and longer follow-up, but require sophisticated methods to account for confounding and censoring when estimating causal treatment effects. A major problem in estimating treatment effects from observational data is confounding by indication; patients receiving more aggressive treatment often have more aggressive disease, creating systematic bias^19^. Standard survival analysis methods such as the Cox proportional hazards model assume constant hazard ratios, precluding assessment of temporal effect dynamics. Causal survival forests, an extension of the generalized random forests framework^20^, enable nonparametric estimation of heterogeneous treatment effects on survival outcomes without imposing proportional hazards assumptions^21,22^. However, these methods produce point estimates at individual time horizons, necessitating methodology to synthesize effects across time into interpretable trajectories.

To address this gap, we apply the Causal Analysis of Survival Trajectories (CAST) framework^23^, a recently developed extension of causal survival forests. CAST was originally introduced and validated in head and neck squamous cell carcinoma and was presented at three NeurIPS 2025 workshops^23^. The method synthesizes horizon-specific treatment effect estimates into smooth temporal curves by estimating the between-horizon covariance structure via bootstrapping and applying Ledoit-Wolf shrinkage^24^ to regularize the resulting covariance matrix. This enables generalized least squares fitting of parametric models alongside nonparametric smoothing splines with bootstrap confidence bands, providing both interpretable summary parameters (peak timing, peak magnitude, half-life) and flexible trajectory characterization. We apply CAST to estimate time-varying effects of radiotherapy and alkylating chemotherapy on overall survival (OS) and progression-free survival (PFS) in two independent cohorts, The Cancer Genome Atlas (TCGA) and the Chinese Glioma Genome Atlas (CGGA), across six treatment-outcome scenarios. By incorporating clinical and molecular covariates (age, sex, WHO grade, IDH mutation status, 1p/19q codeletion, extent of resection) with elastic net propensity score modeling and overlap weighting^25^, and performing subgroup analyses by age, sex, grade, and IDH status, we characterize both average and subgroup-specific temporal treatment dynamics. Treatment effect estimates are validated using three refutation tests and E-value sensitivity analysis^26^. This approach extends prior trial evidence by revealing individualized trajectories of benefit and demonstrates how covariance-aware causal machine learning can move beyond population averages to inform precision treatment strategies.

## Results

### Cohort characteristics

The TCGA cohort comprised up to 512 patients for radiotherapy analyses and 291 patients with detailed chemotherapy annotation for chemotherapy analyses (Supplementary Table S1). The CGGA cohort comprised 264 patients analyzed for both radiotherapy and chemotherapy effects on OS. After six-month landmark analysis to mitigate immortal time bias, sample sizes ranged from 229 to 442 across the six scenarios (Table 1). Kaplan-Meier survival curves by treatment group illustrate the confounding by indication challenge: in several scenarios, treated patients have worse unadjusted survival due to selection of higher-risk patients for treatment (Supplementary Fig. S1). Elastic net propensity score distributions and overlap weight performance are shown in Supplementary Fig. S2.

**Table 1.** Sample sizes after six-month landmark analysis and train/test splits across all six analysis scenarios.

| Scenario | Initial N | Post-landmark N | Train | Test |
| --- | --- | --- | --- | --- |
| TCGA RT OS | 512 | 442 | 332 | 110 |
| TCGA RT PFS | 512 | 423 | 318 | 105 |
| TCGA Chemo OS | 291 | 241 | 181 | 60 |
| TCGA Chemo PFS | 291 | 229 | 172 | 57 |
| CGGA RT OS | 264 | 259 | 195 | 64 |
| CGGA Chemo OS | 264 | 259 | 195 | 64 |

Covariates included age at diagnosis, sex, WHO grade (2 or 3), IDH mutation status, 1p/19q codeletion status, extent of resection (TCGA only, coded categorically), and the alternative treatment modality (e.g., chemotherapy status when analyzing radiotherapy effects). A directed acyclic graph (DAG) was constructed using the dagitty framework^27^ to verify that the selected adjustment set satisfies the backdoor criterion for causal identification. A DAG is a simple diagram where the researcher lays out assumptions about what causes what (e.g., treatment → tumor response → survival, while performance status influences both treatment choice and survival). Evaluating the DAG tells the researcher which variables to adjust for (true confounders) and which to avoid (colliders or mediators), so the estimated treatment effect from observational data is not biased. In short, it helps the researcher make assumptions explicit, choose the right variables, and avoid misleading conclusions from non-randomized oncology data. In this study, our DAG evaluation identified Karnofsky Performance Status (KPS) and tumor location as unmeasured confounders, motivating the comprehensive refutation testing described below.

### Interpreting CAST curves

Before presenting the time-varying treatment effect results, we briefly describe how to read the CAST curve figures. Each CAST curve plots the estimated average treatment effect (ATE) on the y-axis against time horizon on the x-axis. The horizontal dashed line at zero indicates no treatment effect. For survival probability (SP), a positive ATE at time *t* means that treated patients have a higher estimated probability of being alive at *t* months compared to otherwise similar untreated patients; for example, an SP ATE of 0.31 at 84 months indicates an estimated 31 percentage-point higher survival rate at 7 years. For restricted mean survival time (RMST), a positive ATE at time *t* means treated patients are estimated to live that many additional months on average within the first *t* months; for example, an RMST ATE of 18.4 months at 120 months indicates approximately 1.5 years of additional life expectancy within the first decade. Points with error bars show individual horizon-specific estimates with 95% confidence intervals; the solid curve is the nonparametric CAST spline with bootstrap confidence band; the dashed curve is the parametric quadratic CAST fit. Note that y-axes are zoomed to make point estimates clearly visible; error bars that extend beyond the y-axis limits are clipped but not removed.

### Time-varying treatment effects: chemotherapy

Chemotherapy demonstrated consistent positive effects on OS that built progressively over time (Fig. 1A,B). In TCGA, the training-set ATE on survival probability (SP) rose from near zero at 12 months (ATE = -0.02, SE = 0.07) to a sustained plateau at 72-84 months (ATE = 0.30-0.31), before declining at later time points (ATE = 0.14 at 120 months). Test-set predicted ATEs closely tracked training estimates (test ATE = 0.28-0.29 at 72-84 months), indicating good generalization. The parametric CAST model estimated a peak chemotherapy benefit of 0.27 at 76.9 months (approximately 6.4 years post-diagnosis); the nonparametric CAST spline agreed closely (peak 0.24 at 75.3 months). RMST effects were monotonically increasing, reaching 18.4 months gained at 10 years (training ATE; test-set prediction: 16.3 months). E-values peaked at 4.14 (SP at 84 months) and 2.27 (RMST at 120 months), indicating moderate robustness to unmeasured confounding. ATE estimates with 95% confidence intervals at key horizons are reported in Table 2.

**Figure 1.**
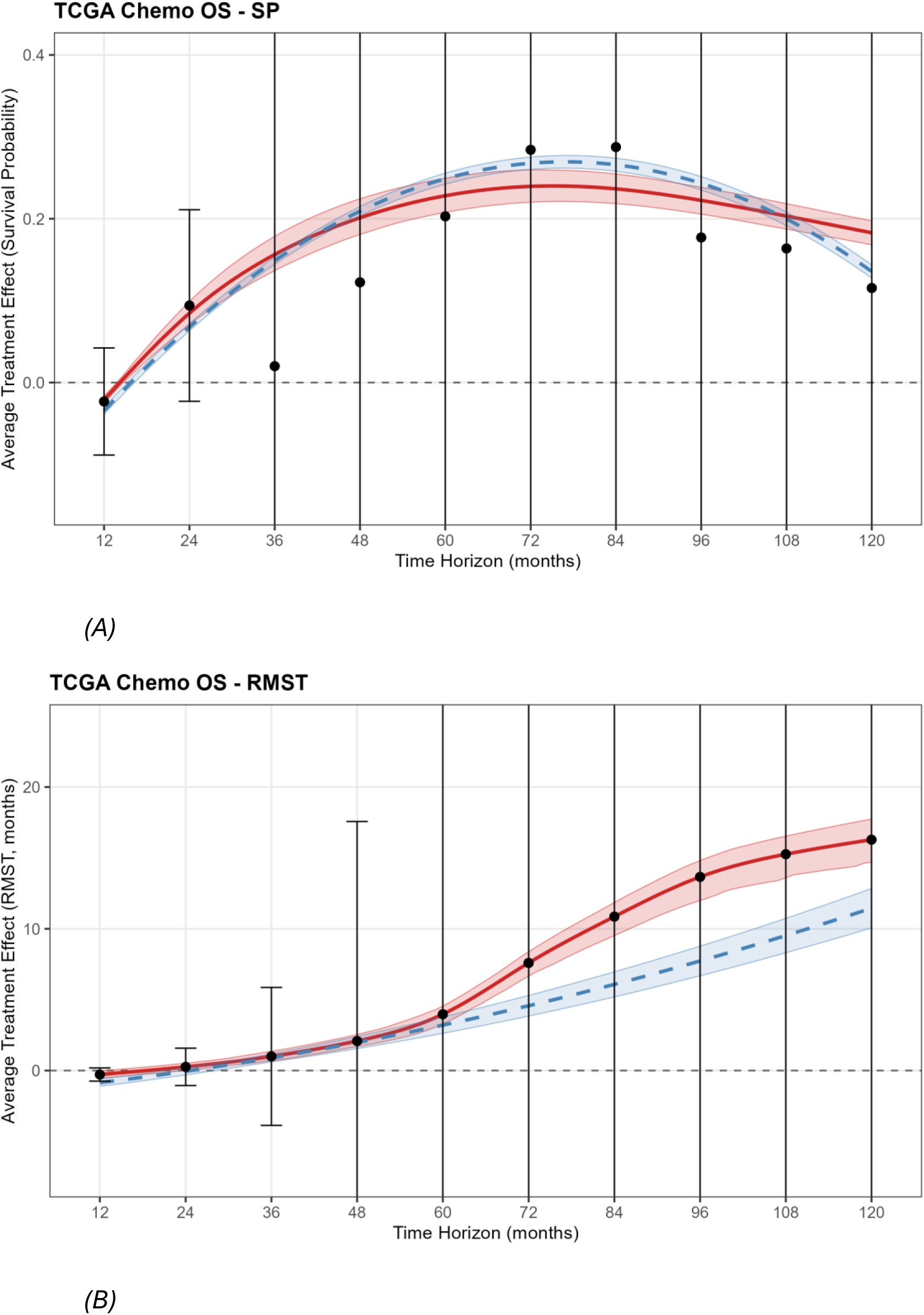
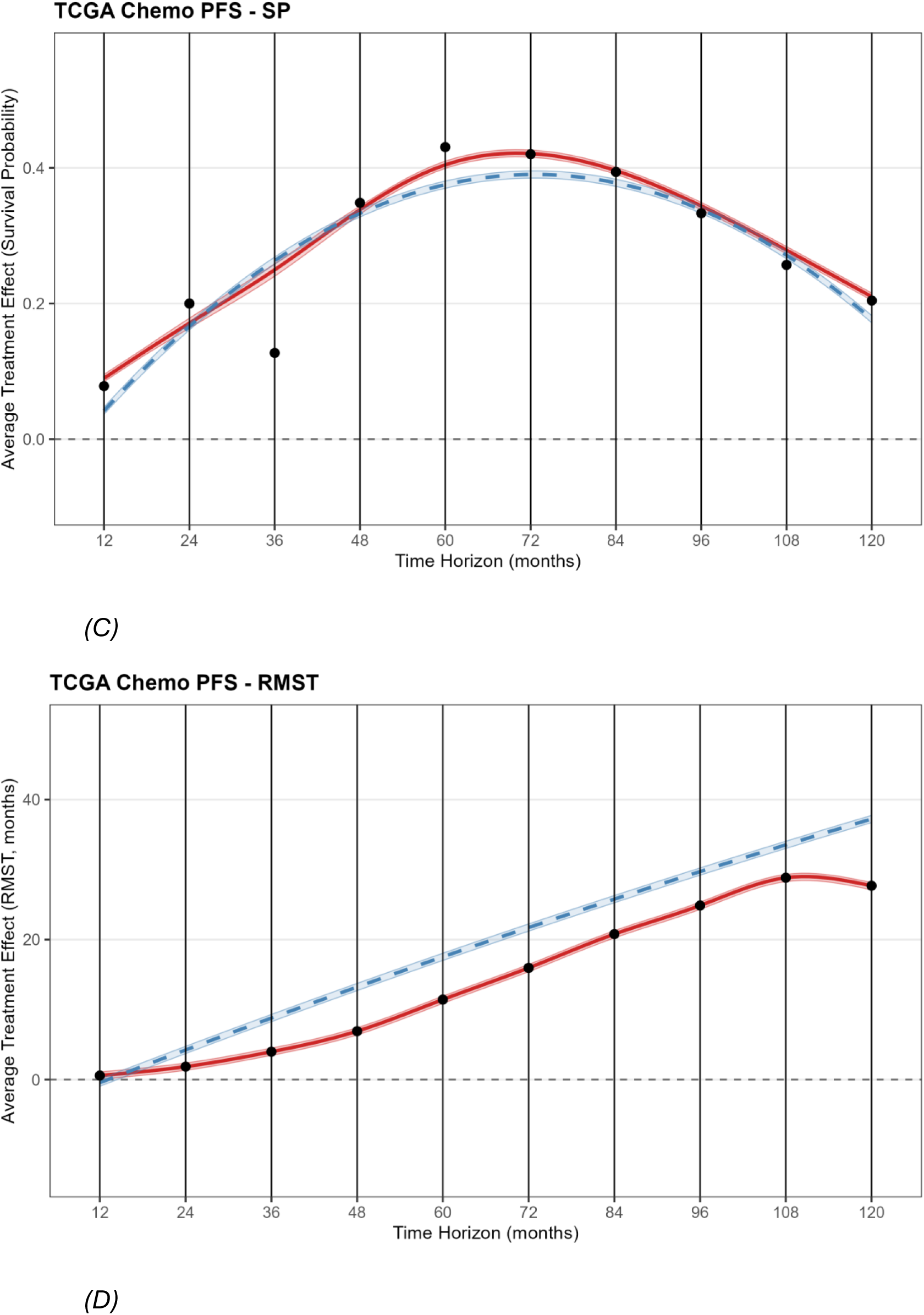
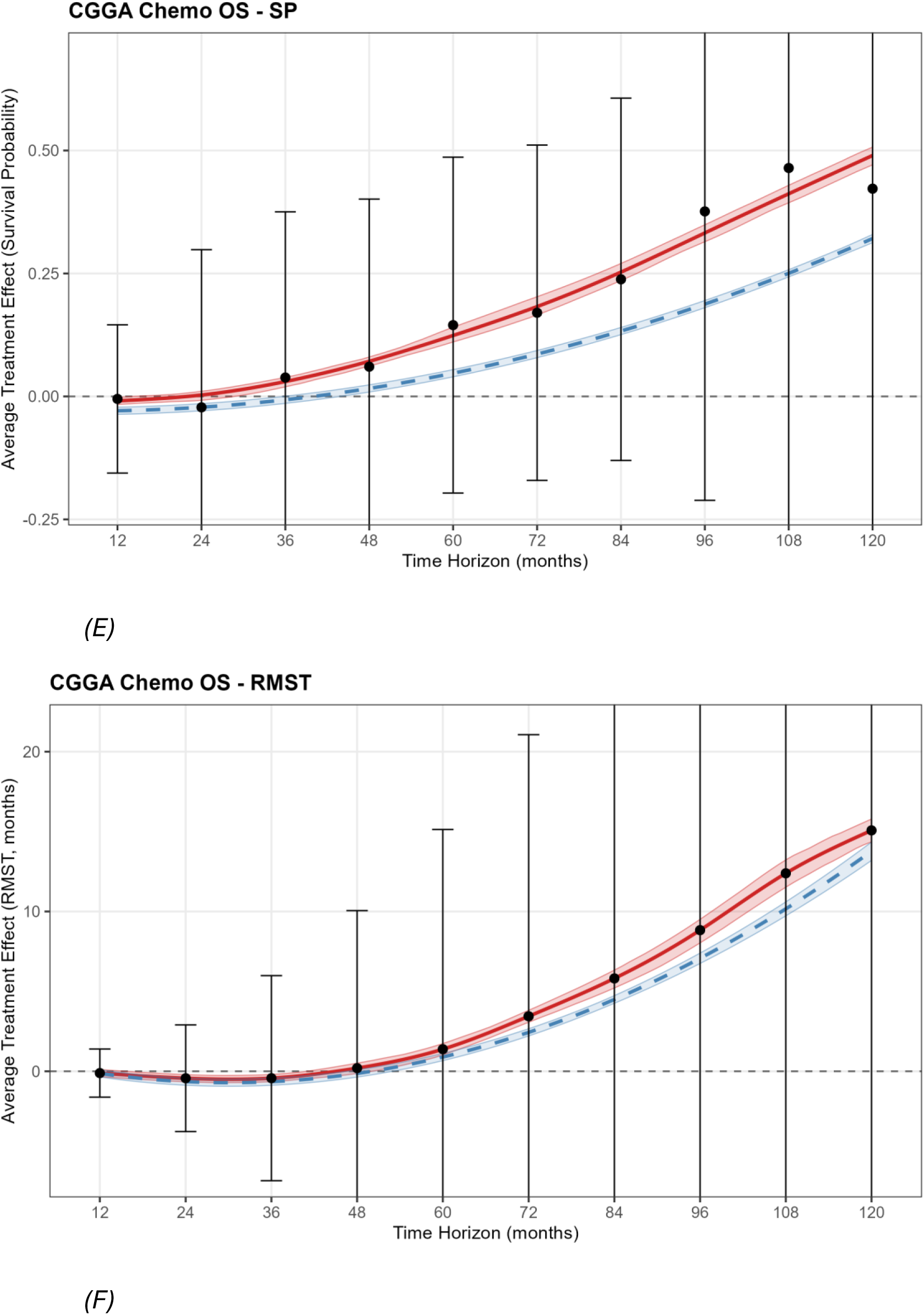
CAST curves for chemotherapy treatment effects. (A,B) TCGA chemotherapy on overall survival: survival probability (SP) and restricted mean survival time (RMST). (C,D) TCGA chemotherapy on progression-free survival: SP and RMST. (E,F) CGGA chemotherapy on overall survival: SP and RMST. Points show horizon-specific ATE estimates with 95% CI; solid curve shows nonparametric CAST spline with bootstrap confidence band; dashed curve shows parametric quadratic CAST fit. The horizontal dashed line at zero indicates no treatment effect. Positive values indicate benefit from chemotherapy. Y-axes are zoomed to emphasize point estimates; error bars extending beyond the plot limits are clipped.

**Table 2.** Average treatment effect (ATE) estimates at selected time horizons.

| Scenario | Horizon (months) | SP ATE (95% CI) | RMST ATE, months<br>(95% CI) | E-value (SP) |
| --- | --- | --- | --- | --- |
| TCGA Chemo OS | 12 | -0.019 (-0.165, 0.126) | -0.268 (-1.337, 0.802) | 1.27 |
| TCGA Chemo OS | 36 | 0.037 (-0.241, 0.314) | 1.374 (-3.442, 6.190) | 1.32 |
| TCGA Chemo OS | 60 | 0.218 (-0.422, 0.857) | 4.978 (-6.900, 16.857) | 2.76 |
| TCGA Chemo OS | 96 | 0.200 (-0.545, 0.945) | 15.325 (-12.942, 43.592) | 2.47 |
| TCGA Chemo PFS | 12 | 0.059 (-0.353, 0.471) | 0.691 (-3.097, 4.478) | 2.53 |
| TCGA Chemo PFS | 36 | 0.125 (-0.438, 0.687) | 3.955 (-8.615, 16.524) | 1.90 |
| TCGA Chemo PFS | 60 | 0.453 (0.022, 0.883) | 13.356 (-3.410, 30.122) | 7.95 |
| TCGA Chemo PFS | 96 | 0.338 (-0.013, 0.689) | 27.688 (2.967, 52.408) | 5.44 |
| TCGA RT OS | 12 | -0.040 (-0.087, 0.008) | -0.147 (-0.326, 0.032) | 2.51 |
| TCGA RT OS | 36 | -0.031 (-0.348, 0.285) | -1.345 (-4.936, 2.246) | 1.78 |
| TCGA RT OS | 60 | 0.162 (-0.430, 0.755) | -0.346 (-10.797, 10.105) | 1.32 |
| TCGA RT OS | 96 | 0.189 (-0.374, 0.752) | 5.454 (-20.702, 31.610) | 3.46 |
| TCGA RT PFS | 12 | 0.068 (-0.169, 0.306) | 0.834 (-0.918, 2.585) | 1.34 |
| TCGA RT PFS | 36 | 0.203 (-0.091, 0.496) | 3.579 (-2.867, 10.024) | 2.30 |
| TCGA RT PFS | 60 | 0.207 (-0.248, 0.662) | 7.053 (-5.161, 19.266) | 2.05 |
| TCGA RT PFS | 96 | 0.204 (-0.219, 0.627) | 14.508 (-8.978, 37.993) | 2.30 |
| CGGA Chemo OS | 12 | -0.005 (-0.110, 0.101) | -0.124 (-0.723, 0.475) | 1.29 |
| CGGA Chemo OS | 36 | 0.028 (-0.133, 0.189) | -0.688 (-3.894, 2.518) | 1.74 |
| CGGA Chemo OS | 60 | 0.177 (-0.039,<br>0.393) | 1.450 (-5.173,<br>8.074) | 5.76 |
| CGGA Chemo OS | 96 | 0.433 (0.087,<br>0.779) | 10.128 (-2.191,<br>22.446) | NA |
| CGGA RT OS | 12 | -0.022 (-0.215,<br>0.171) | 0.212 (-1.565,<br>1.989) | 1.30 |
| CGGA RT OS | 36 | -0.125 (-0.386,<br>0.136) | -1.435 (-8.195,<br>5.325) | 1.78 |
| CGGA RT OS | 60 | -0.232 (-0.514,<br>0.050) | -5.664 (-18.362,<br>7.034) | 3.06 |
| CGGA RT OS | 96 | -0.270 (-0.633,<br>0.093) | -9.263 (-33.424,<br>14.898) | 4.65 |

ATE = average treatment effect; SP = survival probability; RMST = restricted mean survival time; CI = confidence interval. Estimates are from the training set AIPW evaluation. E-values are computed on the held-out test set and quantify robustness to unmeasured confounding; higher values indicate greater robustness.

Chemotherapy effects on PFS were larger than on OS (Fig. 1C,D), with training-set SP ATEs reaching 0.46 at 48 months (test: 0.35) and remaining above 0.40 through 72 months. The parametric CAST peak was 0.39 at 72.6 months (half-life 45.3 months); the nonparametric peak was 0.42 at 70.9 months. RMST effects increased monotonically to 32.5 months at 10 years (training; test: 27.7 months). E-values reached 12.0 (SP at 72 months), indicating strong robustness to unmeasured confounding for the PFS endpoint. The larger PFS effects compared to OS effects are consistent with chemotherapy delaying disease progression as the primary mechanism underlying the OS benefit, aligning with the established efficacy of chemotherapy in lower-grade gliomas from RTOG 9802, EORTC 26951, and CATNON^10–13^.

In CGGA, chemotherapy demonstrated delayed positive effects on OS (Fig. 1E,F). SP ATEs were near zero at 12-36 months, then rose to 0.18 at 60 months, 0.28 at 84 months, and peaked at 0.48 at 108 months before declining to 0.44 at 120 months. Test-set predictions closely tracked training estimates (test ATE = 0.46 at 108 months). RMST effects were initially negative but turned positive by 60 months, reaching 16.8 months at 10 years (training; test: 15.1 months). E-values were exceptionally large at later horizons (27.6 at 108 months for SP), indicating that the late chemotherapy benefit is highly robust to unmeasured confounding. The delayed emergence of benefit in CGGA relative to TCGA (108 vs. 72-84 months for peak SP effect) may reflect differences in patient populations, molecular subtype distributions, chemotherapy regimens, or healthcare systems between Western and Chinese glioma cohorts^28,29^.

### Time-varying treatment effects: radiotherapy

Radiotherapy effects on OS exhibited a transition from negative effects at early horizons to modest positive effects at later horizons in TCGA (Fig. 2A,B). SP ATEs were negative from 12 to 48 months (training ATE = -0.08 at 24 months, -0.06 at 48 months), became positive at 60 months (ATE = 0.16), and peaked at 72 and 96 months (ATE = 0.19), with a slight decrease at 84 months (ATE = 0.14). Test-set predictions showed a similar transition with attenuated magnitudes. RMST effects followed the same pattern: negative through 60 months (training: -1.7 months at 48 months), turning positive at 72 months and reaching +8.7 months at 120 months (training; test: +2.2 months). E-values for the RT OS effects were modest (1.3-4.6 for SP), indicating that the observed effects are sensitive to unmeasured confounding. The initial negative effects likely reflect confounding by indication (patients receiving radiotherapy had more aggressive disease features) while the late positive effects may represent a delayed survival benefit emerging after the confounding signal attenuates.

**Figure 2.**
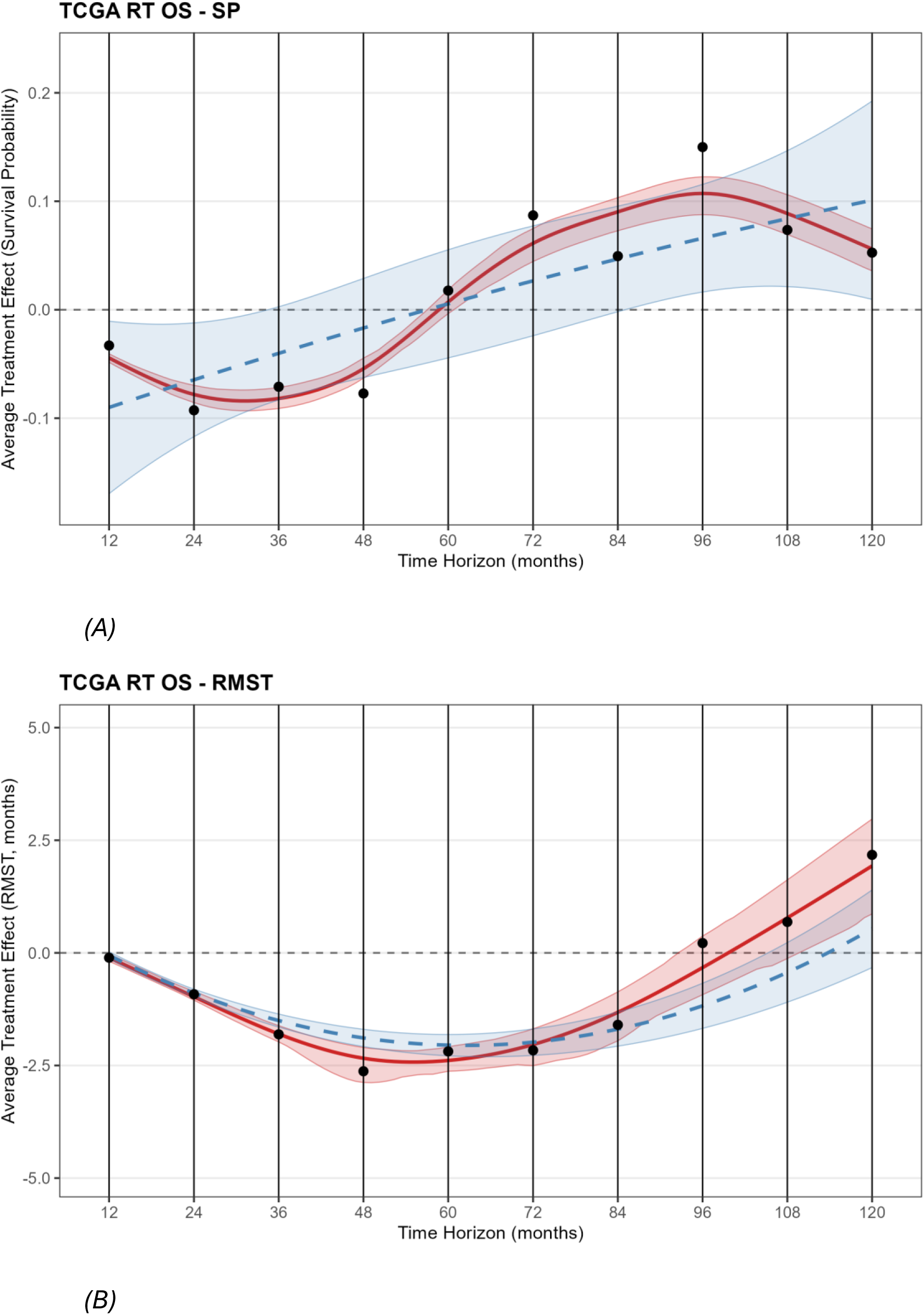
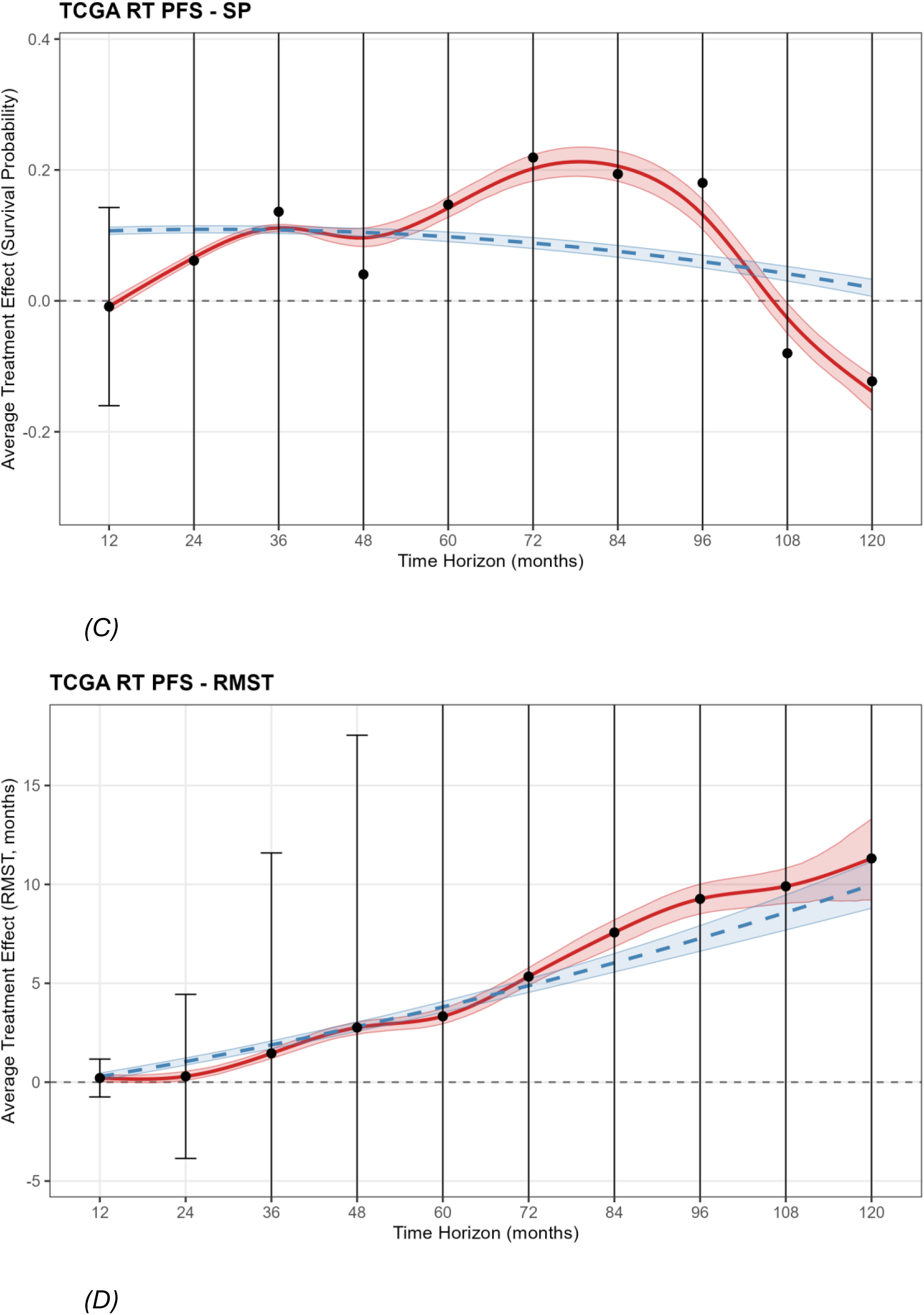
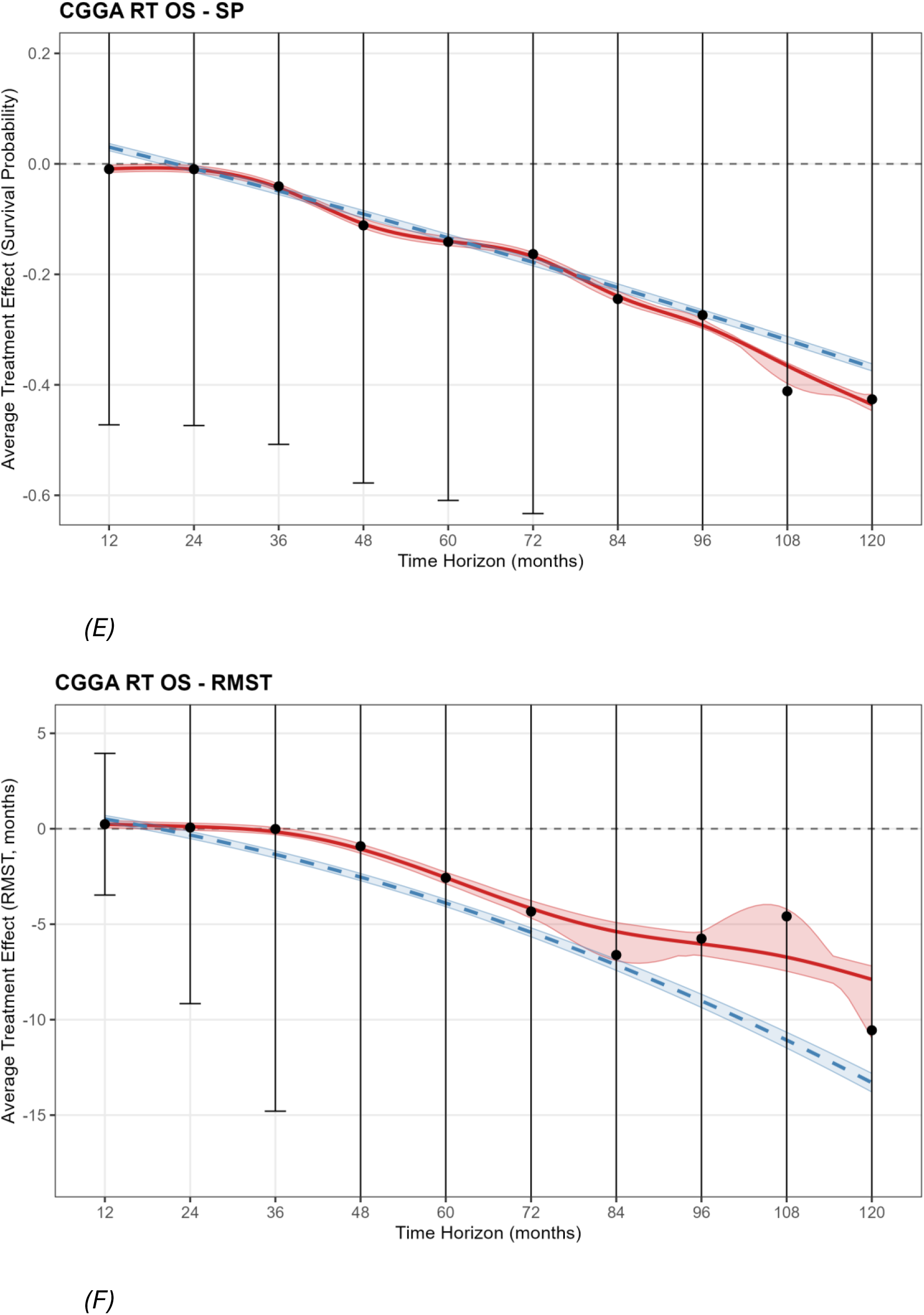
CAST curves for radiotherapy treatment effects. (A,B) TCGA radiotherapy on overall survival: SP and RMST. (C,D) TCGA radiotherapy on progression-free survival: SP and RMST. (E,F) CGGA radiotherapy on overall survival: SP and RMST. In TCGA OS, the CAST curve transitions from negative at early horizons (reflecting confounding by indication) to modestly positive at later horizons. TCGA PFS shows generally positive effects. CGGA RT shows consistently negative effects, likely reflecting stronger residual confounding due to absence of extent of resection data. Y-axes are zoomed; error bars extending beyond the plot limits are clipped.

Radiotherapy effects on PFS were more favorable than on OS, with positive effects throughout most of the follow-up period (Fig. 2C,D). SP ATEs built from 0.07 at 12 months to a peak of 0.25 at 72 months (training), before declining and reversing at 108-120 months. The nonparametric CAST peak was 0.21 at 79.6 months. RMST effects were consistently positive and monotonically increasing, reaching 16.9 months at 10 years (training; test: 11.3 months). The more favorable PFS profile compared to OS suggests that radiotherapy provides local tumor control benefit that is partially obscured in OS analyses by residual confounding from unmeasured disease severity factors.

In CGGA, radiotherapy effects were consistently negative across all horizons (Fig. 2E,F). SP ATEs worsened progressively from -0.02 at 12 months to -0.37 at 84 months and -0.75 at 120 months (training). E-values for the negative RT effect were moderate (1.3-4.7). These persistently negative effects likely reflect confounding by indication that is not fully resolved by the available covariates. The absence of extent of resection data in CGGA, a known confounder of both treatment decisions and outcomes^30^, likely contributes to stronger residual confounding compared to TCGA.

### Subgroup heterogeneity

Subgroup-specific CAST curves were generated for age (above/below median), sex, WHO grade, and IDH mutation status (Fig. 3). These reveal how the temporal treatment effect trajectory varies across patient subgroups.

**Figure 3.**
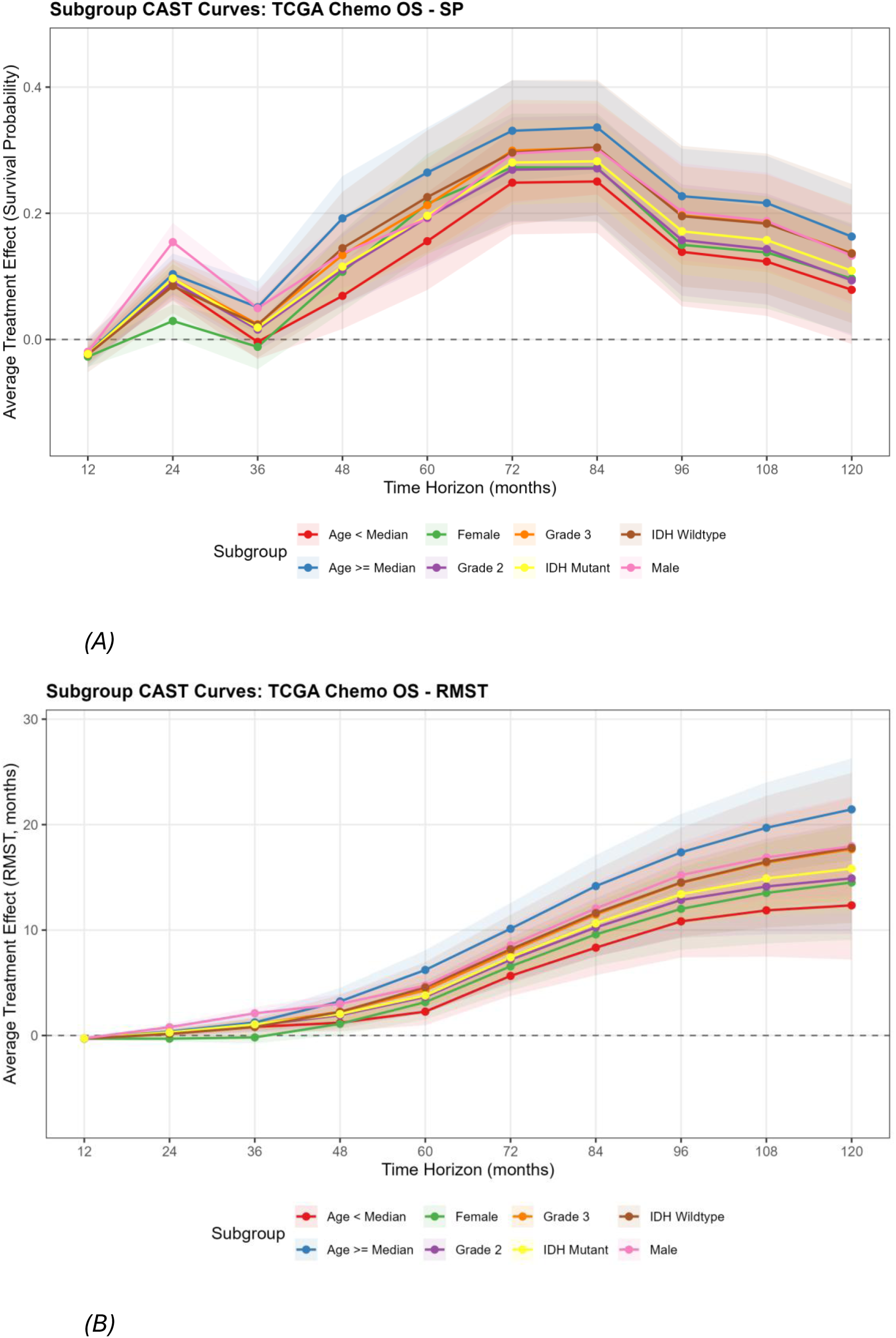
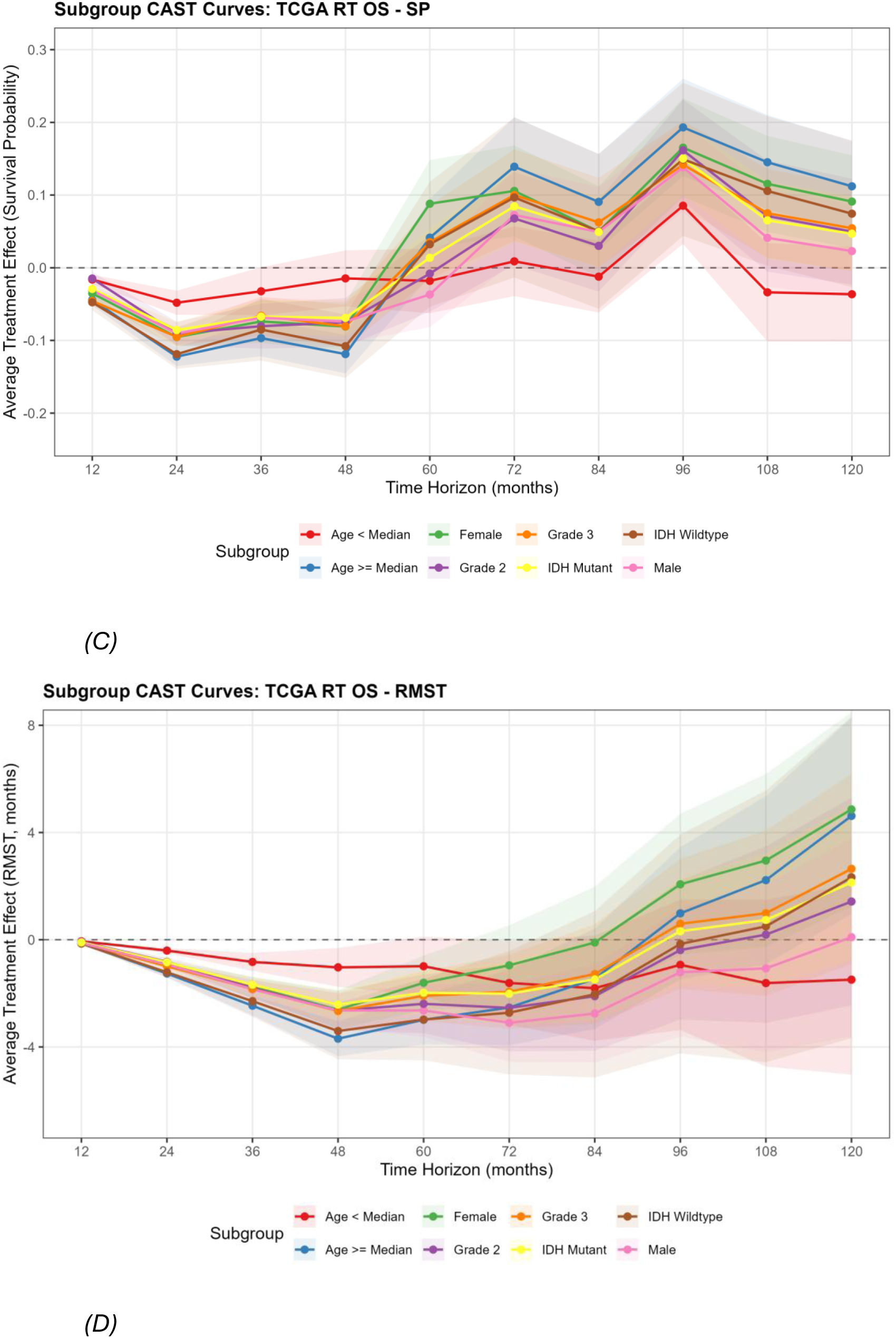
Subgroup CAST curves for TCGA overall survival. (A,B) Chemotherapy subgroups: SP and RMST. (C,D) Radiotherapy subgroups: SP and RMST. Subgroup-specific trajectories are shown for age groups (above/below median), sex, WHO grade, and IDH mutation status. All subgroups show positive chemotherapy effects at later horizons, with effect heterogeneity driven primarily by age. Radiotherapy subgroups show the characteristic early negative-to-late positive transition, with older patients demonstrating larger late benefits. Y-axes are zoomed; error bars extending beyond the plot limits are clipped.

For TCGA chemotherapy on OS (Fig. 3A,B), older patients (age at or above the median) showed larger treatment effects than younger patients at most horizons (RMST ATE at 84 months: 14.2 vs. 8.3 months), consistent with the dominant role of age in the variable importance analysis. Male patients showed modestly larger effects than females (12.1 vs. 9.6 months at 84 months). Grade 3 patients showed larger effects than grade 2 patients (11.5 vs. 10.3 months at 84 months). IDH-mutant and IDH-wildtype subgroups showed similar effect magnitudes (10.6 vs. 11.6 months at 84 months), suggesting that while IDH status is strongly prognostic, it is not a major driver of chemotherapy *treatment effect* heterogeneity in this analysis.

For TCGA radiotherapy on OS (Fig. 3C,D), older patients again showed larger positive effects at later horizons, while all subgroups exhibited the early negative-to-late positive transition reflecting confounding. CGGA chemotherapy subgroup effects followed similar patterns but with the delayed temporal profile characteristic of the CGGA cohort: older patients showed larger effects at later horizons, and IDH-mutant patients showed stronger late effects (RMST ATE at 120 months: 15.9 vs. 12.4 months for IDH-wildtype). CGGA radiotherapy showed consistently negative effects across all subgroups, with older patients and IDH-wildtype patients exhibiting the most negative effects.

### Feature importance and cross-cohort comparison

Variable importance analysis from the causal survival forests identified age at diagnosis as the dominant driver of treatment effect heterogeneity across all six scenarios, accounting for 47-56% of splits (Supplementary Table S2). In TCGA, extent of resection was the second most important feature (13-19%), followed by sex (15-18%) and WHO grade (8-12%). In CGGA, which lacks EOR data, grade (15-16%) and 1p/19q codeletion (13%) emerged as the second and third most important features, likely absorbing variance that EOR captures in TCGA. IDH mutation status showed zero importance across all scenarios despite its well-known prognostic significance, reflecting the distinction between prognostic and predictive biomarkers: after including 1p/19q codeletion and grade, IDH provides no additional information for partitioning treatment effect heterogeneity.

CAST curve parameters are summarized in Supplementary Table S3. Chemotherapy effects were consistently positive in both cohorts, with TCGA showing earlier benefit emergence (24-36 months) and CGGA showing delayed but comparable magnitudes. Radiotherapy effects were more negative in CGGA than in TCGA, likely reflecting stronger residual confounding due to the absence of extent of resection data.

### Validation and refutation tests

Three classes of refutation tests assessed the robustness of causal effect estimates (Supplementary Figs. S3-S4). Placebo (treatment permutation) tests produced ATE estimates centered around zero across all horizons and metrics, confirming that the causal forest does not generate spurious effects when the treatment-outcome relationship is destroyed. Mean absolute ATEs under permutation were consistently below 0.05 for SP and below 1.0 month for RMST across all 20 replicates per scenario. Unobserved confounder sensitivity tests introduced synthetic confounders at three strength levels (gamma = 0.1, 0.3, 0.5). At low and moderate strengths, ATE perturbations generally remained within standard error bounds; at high strength, meaningful shifts were observed at longer horizons where censoring reduces effective sample size. Negative control tests confirmed null ATEs under random treatment assignment and stability of ATEs when five Gaussian noise covariates were added alongside the real treatment.

## Discussion

Our analysis of 776 patients from TCGA and CGGA reveals distinct temporal treatment effect patterns with implications for precision oncology. The CAST framework, applied here to lower-grade gliomas, enables visualization of how treatment benefits emerge, peak, and evolve over time, moving beyond the static hazard ratios of traditional survival analysis and the isolated time-point estimates of standard causal survival forests. By synthesizing multiple horizon-specific effect estimates into coherent temporal trajectories with proper uncertainty quantification, CAST provides a principled approach for characterizing treatment effect dynamics.

The central finding is the consistent positive chemotherapy effect across both cohorts and across both OS and PFS endpoints. In TCGA, chemotherapy effects on survival probability peaked at 0.31 at 72-84 months for OS (an estimated 31 percentage-point higher chance of being alive at 7 years) and 0.46 at 48 months for PFS, with RMST gains of 18.4 and 32.5 months at 10 years, respectively. In CGGA, chemotherapy showed delayed positive effects on OS, peaking at 0.48 at 108 months with RMST gains of 16.8 months at 10 years. These findings corroborate landmark trials (RTOG 9802, RTOG 9402/EORTC 26951, CATNON) while revealing temporal dynamics obscured by fixed-interval reporting^10–13^. The larger PFS effects compared to OS effects in TCGA are biologically plausible, suggesting that treatment delays disease progression as the primary mechanism underlying the OS benefit. The delayed emergence of chemotherapy benefit in CGGA relative to TCGA (108 vs. 72-84 months for peak SP effect) may reflect differences in patient populations, molecular subtype distributions, or healthcare systems between Western and Chinese glioma cohorts^28,29,31^. By analyzing diverse international cohorts separately, we demonstrate that findings can replicate across distinct populations while preserving cohort-specific characteristics.

Radiotherapy effects were more complex and differed between cohorts. In TCGA, radiotherapy showed negative effects at early horizons transitioning to modest positive effects later for OS, and generally positive effects on PFS. In CGGA, radiotherapy effects were consistently negative across all horizons. Several factors likely explain this pattern. First, confounding by indication is expected to be stronger for radiotherapy than chemotherapy; radiotherapy is more commonly administered to patients with aggressive disease features (higher grade, less complete resection, IDH-wildtype status), as confirmed by the strong propensity score predictors in our RT analyses. Second, the absence of extent of resection data in CGGA likely amplifies residual confounding, since surgical extent influences both RT decisions and outcomes^30^. Third, the modest E-values for RT effects (1.1-4.7) indicate sensitivity to unmeasured confounding. Performance status (KPS), a strong prognostic factor and major treatment selection criterion unavailable in either dataset, may contribute substantially to residual confounding, particularly for radiotherapy analyses. The positive TCGA RT PFS effects (peak SP 0.25 at 72 months; RMST 16.9 months at 10 years), combined with the late positive RT OS effects, suggest a genuine treatment benefit that is partially masked by confounding in OS analyses, consistent with the established efficacy of RT in lower-grade gliomas, particularly on PFS, from randomized trials^10,32,33^.

The CAST framework provides a principled approach for visualizing temporal treatment effect dynamics. Causal survival forests estimate separate treatment effects at each time horizon, but these estimates are correlated because they arise from the same patients. CAST addresses this by estimating the between-horizon covariance structure via bootstrapping and applying Ledoit-Wolf shrinkage^24^ for regularization. The combination of parametric and nonparametric CAST models offers complementary perspectives: the quadratic parametric model provides interpretable summary statistics (peak timing, peak magnitude, half-life) while the smoothing spline captures complex trajectory shapes without parametric assumptions. This covariance-aware approach^23^ accounts for patient-level dependencies that are ignored by naive time-point-by-time-point analyses.

Rather than acting uniformly over the disease course, radiotherapy and chemotherapy exert their strongest effects at different times: chemotherapy appears to provide earlier disease control, while radiotherapy may offer more durable long-term survival benefits. This temporal distinction supports treatment strategies that balance short-term stabilization against long-term outcomes.

Variable importance analysis consistently identified age at diagnosis as the dominant treatment effect heterogeneity driver (46-56% of splits), aligning with clinical knowledge that treatment response varies with age^34^. The prominence of extent of resection in TCGA (13-19%) underscores the interaction between surgical completeness and adjuvant treatment efficacy^30^. In CGGA, 1p/19q codeletion status emerged as a key heterogeneity driver (13%), likely compensating for the absence of EOR data. The zero importance of IDH mutation status reflects its high correlation with 1p/19q codeletion and grade rather than a lack of biological relevance, highlighting the important distinction between prognostic and predictive biomarkers.

### Limitations

This retrospective analysis has several limitations. Potential unmeasured confounding remains a concern despite refutation tests that support robustness. Several important prognostic variables were unavailable in the TCGA and CGGA datasets, including performance status, MGMT promoter methylation status, specific chemotherapy regimen (temozolomide vs. PCV could not be distinguished), radiotherapy dose and fractionation schedules, and tumor size. However, in lower-grade gliomas, most patients have high performance status (KPS >70), potentially reducing the magnitude of this confounding. The DAG evaluation confirmed that the available adjustment set does not fully block all backdoor paths due to unmeasured confounders (KPS, tumor location). The TCGA chemotherapy analyses used a 291-patient annotated subset where the propensity model found no measured confounders of treatment assignment; while this simplifies interpretation, unmeasured treatment selection factors remain unadjusted. CGGA lacked PFS outcomes and EOR data, limiting cross-cohort comparisons. Censoring at long horizons (96-120 months) reduces statistical power and may introduce bias. We did not adjust for multiple comparisons across time horizons, instead relying on CAST trajectory modeling to assess overall temporal patterns. The six-month landmark analysis, while mitigating immortal time bias, excludes early events that may be treatment-related. Limited long-term follow-up beyond 10 years may miss very late treatment effects or toxicities.

## Conclusions

By quantifying when treatment benefits emerge and attenuate, our results provide a quantitative foundation for individualized treatment planning that considers both patient characteristics and the timing of therapeutic benefit. We apply the CAST framework, a recently developed extension of causal survival forests that synthesizes horizon-specific causal treatment effect estimates into smooth temporal trajectories while accounting for between-horizon covariances. Applied to lower-grade gliomas, CAST curves reveal that chemotherapy demonstrates consistent positive effects across both TCGA and CGGA cohorts, with survival probability gains of 0.31-0.48 and RMST gains of 16.8-32.5 months, concordant with landmark RCT evidence. Radiotherapy effects are mixed, with modest E-values indicating sensitivity to residual confounding by indication rather than true harm. The CAST framework, integrating Ledoit-Wolf covariance regularization, parametric peak characterization, and nonparametric confidence bands, provides a general-purpose tool for temporal treatment effect visualization applicable across clinical domains. Future work should validate these temporal patterns in prospective cohorts with standardized molecular characterization and longer follow-up.

## Methods

### Study populations

We used the CAST (Causal Analysis of Survival Trajectories) framework^23^ to estimate time-varying effects of radiotherapy and alkylating chemotherapy on survival in adults with WHO grade 2-3 diffuse gliomas. Two publicly available datasets were analyzed independently. The Cancer Genome Atlas (TCGA) included adult diffuse glioma patients with clinical, molecular, and treatment data^6^. For radiotherapy analyses, the full TCGA dataset (512 patients across multiple institutions) was used. For chemotherapy analyses, a 291-patient subset with detailed chemotherapy annotation was used, where chemotherapy status codes were binarized (no chemotherapy vs. any chemotherapy). The Chinese Glioma Genome Atlas (CGGA) included 264 adult diffuse glioma patients from two institutional datasets (CGGA-325 and CGGA-693 batches combined)^31^. Only OS outcomes were available in CGGA. Both cohorts included patients with WHO grade 2-3 diffuse gliomas. Separate analysis of the two cohorts avoided dataset-specific biases, ensured consistent covariate availability, and enabled replication across distinct populations.

### Causal framework and covariate selection

A directed acyclic graph (DAG) was constructed using the dagitty framework^27^ to formalize assumptions about the causal structure and verify that the adjustment set satisfies the backdoor criterion. Covariates included age at diagnosis (continuous), sex (binary), WHO grade (2 or 3), IDH mutation status (mutant vs. wildtype), 1p/19q codeletion status, extent of resection (TCGA only, categorical), and the alternative treatment modality (e.g., chemotherapy status when analyzing radiotherapy effects). EOR was unavailable in CGGA and was excluded from CGGA analyses. Variables with >25% missingness were excluded prior to imputation (e.g., chemotherapy status had 43.3% missingness in TCGA RT analyses).

### Data preprocessing

A six-month landmark analysis was applied to mitigate immortal time bias^35^, excluding patients who died or were censored within six months and adjusting survival times accordingly. Missing values were imputed using a cascaded approach: missForest^36^ (random forest-based imputation excluding outcome and treatment variables) was attempted first, with fallback to multiple imputation by chained equations (MICE; m = 20 imputations combined using simplified Rubin’s rules)^37^, and finally median/mode imputation. Data were split into training (75%) and test (25%) sets using stratified sampling to preserve the treatment-outcome distribution.

### Propensity score estimation and overlap weighting

Treatment propensity scores were estimated using elastic net logistic regression^38^ with the mixing parameter alpha selected via cross-validation over [0.01, 0.99] and lambda chosen to minimize cross-validated deviance. Rather than trimming patients with extreme propensity scores, we employed overlap weighting^25^: treated patients received weight (1 - e(X)), untreated patients received weight e(X), where e(X) is the estimated propensity score. Overlap weights target the average treatment effect in the overlap population, upweighting patients in the region of clinical equipoise where treated and untreated groups are most comparable. Balance was assessed using standardized mean differences and variance ratios.

### Causal survival forests

Causal survival forests from the grf package^20,21^ estimated conditional average treatment effects (CATEs) on two survival metrics at each of 10 time horizons (12, 24, 36, 48, 60, 72, 84, 96, 108, 120 months post-diagnosis): survival probability (SP) differences and restricted mean survival time (RMST) differences. Each forest used 5,000 trees with propensity scores supplied and overlap weights via the sample.weights parameter. The ATE at each horizon was the weighted mean of individual CATEs. E-values were computed following VanderWeele and Ding^26^ to quantify robustness to unmeasured confounding. Variable importance was extracted via the grf variable_importance() function and aggregated across horizons.

### CAST curve methodology

To synthesize horizon-specific ATE estimates into temporal trajectories, we applied the CAST framework^23^, which was originally developed for head and neck squamous cell carcinoma and presented at three NeurIPS 2025 workshops (AI4Science, CauScien, TS4H). ATE estimates at neighboring horizons are correlated because they arise from the same patients. We estimated the between-horizon covariance via bootstrapping test-set predictions (200 replicates) and applied Ledoit-Wolf shrinkage^24^ to the resulting covariance matrix:

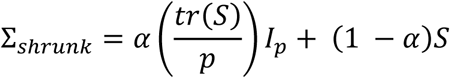

where S is the bootstrap sample covariance, p is the dimension, and alpha is the data-driven optimal shrinkage intensity.

A quadratic parametric model was fitted via generalized least squares (GLS) using the shrinkage-regularized covariance, with fallback to weighted least squares:

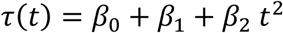

The parametric model provided estimates of peak timing 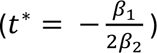, peak effect magnitude, and effect half-life. A nonparametric smoothing spline weighted by inverse standard errors, with 95% confidence bands from parametric bootstrap (200 replicates), captured flexible treatment trajectories without assuming a specific functional form.

### Subgroup analyses

Subgroup-specific CAST curves were generated for age groups (above/below median), sex, WHO grade, and IDH mutation status. For each subgroup, patients were filtered to the relevant subgroup and weighted averages of individual CATEs were computed using overlap weights. Subgroup CAST curves with 95% confidence intervals were generated to visualize treatment effect variation across subgroups and over time.

### Refutation and validation tests

Three classes of refutation tests assessed the robustness of causal effect estimates. Placebo tests randomly permuted the treatment vector (20 replicates per horizon) to verify null ATEs under no treatment-outcome association. Unobserved confounder sensitivity tests introduced a synthetic confounder correlated with age at strengths gamma = 0.1, 0.3, and 0.5, modifying both treatment assignment and outcomes, then refitting the forest without the confounder and measuring ATE distortion (20 replicates per strength). Negative control tests verified null ATEs under random treatment assignment and stability of ATEs when five Gaussian noise covariates were added alongside the real treatment.

### Statistics and reproducibility

All analyses were conducted in R version 4.5.1. Key packages included grf (v2.5.0) for causal survival forests, glmnet (v4.1.10) for elastic net propensity models, survival (v3.8.3) and survminer for Kaplan-Meier analyses, missForest and mice for imputation, and dagitty and ggdag for DAG evaluation. All statistical tests were two-sided. Sample sizes were determined by the available data in each publicly accessible dataset; no a priori power calculations were performed. Results were replicated independently in TCGA and CGGA cohorts to assess robustness across populations.

## Ethics statement

The TCGA and CGGA datasets are publicly available and contain fully de-identified patient data. All original studies obtained institutional review board approval and informed consent. No additional ethics approval was required for this secondary analysis.

## Data availability

The TCGA lower-grade glioma and glioblastoma datasets are publicly available through the Genomic Data Commons (GDC) Data Portal (https://portal.gdc.cancer.gov/). The CGGA dataset is publicly available through the Chinese Glioma Genome Atlas website (http://www.cgga.org.cn/).

## Code availability

All analysis code is available at https://github.com/everestyang1/final-lgg-cast. The CAST framework code is available at https://github.com/CAST-FW/HNSCC.

## Acknowledgments

There was no outside funding for this project.

## Author contributions

E.Y and I.S. conceived the study, designed and implemented the causal inference pipeline, including the CAST curve methodology, causal survival forest analyses, propensity score modeling, refutation tests, and DAG evaluation framework. E.Y. developed the initial CAST methodology, contributed to data curation and harmonization, analysis implementation, and drafted the manuscript. S.A. contributed to analysis support, and biological interpretation. C.J.K. provided neuro-oncology clinical domain expertise, critical review of results interpretation, and guidance on covariate selection. L.Y. and E.W. contributed to data curation and analysis support. S.K.C, T.J.W. and L.A.K. provided clinical radiation oncology expertise and critical review of clinical interpretations. D.J.B. contributed to study design, methodological guidance, and interpretation. All authors reviewed, edited, and approved the final manuscript.

## Supplementary Materials

**Supplementary Table S1:** Patient Characteristics.

| Characteristic | Overall | RT | No RT | Chemo | No Chemo |
| --- | --- | --- | --- | --- | --- |
| Age, years, mean (SD) | 42.6 (13.5) | 43.3 (13.9) | 41.2 (12.7) | 43.3 (13.8) | 39.9 (12.4) |
| Male sex, n (%) | 162 (55.7%) | 117 (58.5%) | 45 (49.5%) | 131 (56.5%) | 31 (52.5%) |
| WHO Grade 2, n (%) | 143 (49.1%) | 74 (37.0%) | 69 (75.8%) | 107 (46.1%) | 36 (61.0%) |
| WHO Grade 3, n (%) | 147 (50.5%) | 125 (62.5%) | 22 (24.2%) | 124 (53.4%) | 23 (39.0%) |
| IDH-mutant, n (%) | 229 (78.7%) | 150 (75.0%) | 79 (86.8%) | 178 (76.7%) | 51 (86.4%) |
| IDH-wildtype, n (%) | 56 (19.2%) | 46 (23.0%) | 10 (11.0%) | 49 (21.1%) | 7 (11.9%) |
| 1p/19q codeletion, n (%) | 229 (78.7%) | 150 (75.0%) | 79 (86.8%) | 178 (76.7%) | 51 (86.4%) |
| OS, months, median (IQR) | 22.1 (9.9-42.0) | 22.9 (13.2-42.6) | 20.6 (5.7-39.8) | 19.6 (7.4-40.2) | 27.5 (18.6-46.2) |
| Deaths, n (%) | 74 (25.4%) | 62 (31.0%) | 12 (13.2%) | 51 (22.0%) | 23 (39.0%) |
| PFS, months, median (IQR) | 17.3 (6.5-35.6) | 17.5 (6.8-37.9) | 16.5 (4.2-30.1) | 16.6 (6.3-35.4) | 20.7 (12.6-36.0) |
| PFS events, n (%) | 108 (37.1%) | 79 (39.5%) | 29 (31.9%) | 77 (33.2%) | 31 (52.5%) |

CGGA Cohort
| Characteristic | Overall | RT | No RT | Chemo | No Chemo |
| --- | --- | --- | --- | --- | --- |
| Age, years, mean (SD) | 39.9 (10.5) | 41.0 (10.2) | 36.5 (10.9) | 40.7 (10.2) | 38.4 (11.0) |
| Male sex, n (%) | 147 (55.7%) | 107 (54.3%) | 40 (59.7%) | 96 (56.5%) | 51 (54.3%) |
| WHO Grade 2, n (%) | 127 (48.1%) | 90 (45.7%) | 37 (55.2%) | 67 (39.4%) | 60 (63.8%) |
| WHO Grade 3, n (%) | 137 (51.9%) | 107 (54.3%) | 30 (44.8%) | 103 (60.6%) | 34 (36.2%) |
| IDH-mutant, n (%) | 175 (74.8%) | 133 (76.0%) | 42 (71.2%) | 113 (74.3%) | 62 (75.6%) |
| IDH-wildtype, n (%) | 59 (25.2%) | 42 (24.0%) | 17 (28.8%) | 39 (25.7%) | 20 (24.4%) |
| 1p/19q codeletion, n (%) | 168 (63.6%) | 127 (64.5%) | 41 (61.2%) | 109 (64.1%) | 59 (62.8%) |
| OS, months, median (IQR) | 56.2 (33.2-83.9) | 55.7 (36.2-79.9) | 65.8 (29.9-91.8) | 64.5 (33.8-86.4) | 53.1 (32.4-74.1) |
| Deaths, n (%) | 95 (36.0%) | 79 (40.1%) | 16 (23.9%) | 59 (34.7%) | 36 (38.3%) |

N = 264 patients from CGGA LGG cohort. PFS data and EOR data not available in the CGGA cohort. Follow-up times represent the median observed time across all patients (censored and uncensored), not Kaplan-Meier estimated median survival.

Supplementary Table S2: Feature Importance

**Supplementary Table S2.**
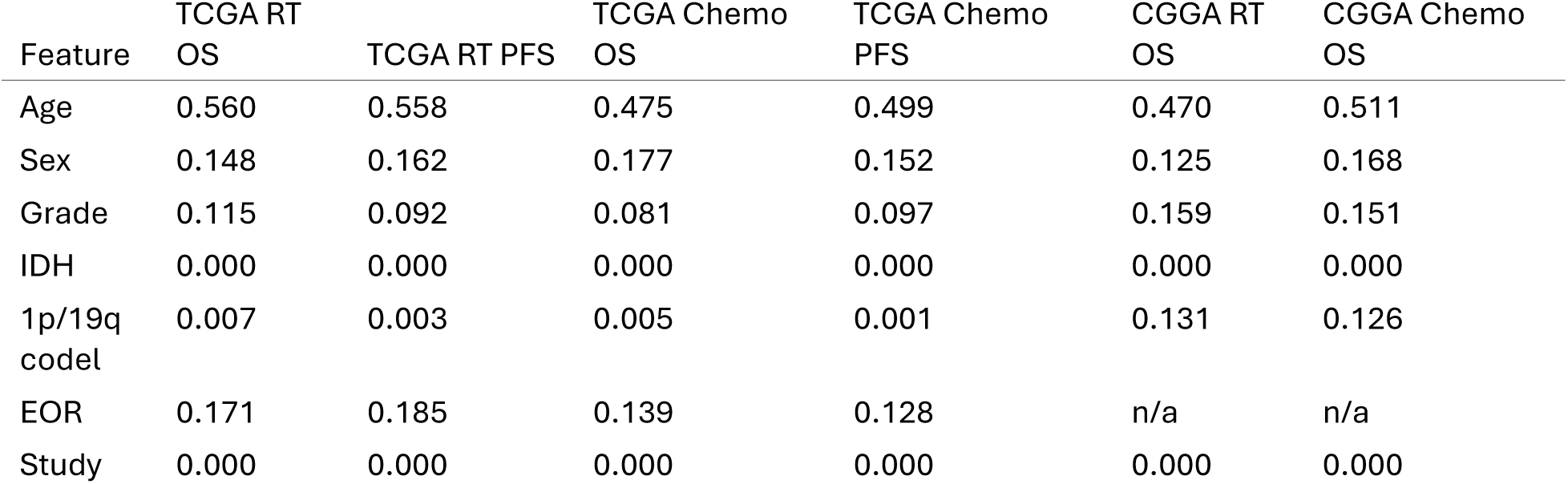
Mean variable importance (proportion of splits) for the survival probability metric, averaged across all time horizons.

Supplementary Table S3: CAST Curve Parameters

**Supplementary Table S3.** Summary of CAST curve parameters across all analysis scenarios.

| Scenario | Metric | Parametric peak | Peak time (months) | Half-life (months) | NP peak | NP peak time (months) |
| --- | --- | --- | --- | --- | --- | --- |
| TCGA Chem o OS | SP | 0.270 | 76.9 | 43.2 | 0.240 | 75.3 |
| TCGA Chem o PFS | SP | 0.390 | 72.6 | 45.3 | 0.422 | 70.9 |
| TCGA RT OS | SP | n/a | n/a | n/a | 0.107 | 97.1 |
| TCGA RT PFS | SP | 0.109 | 26.7 | 72.9 | 0.212 | 79.6 |
| CGGA Chem o OS | SP | -0.030 | 7.2 | 23.2 | n/a | n/a |
| CGGA RT OS | SP | n/a | n/a | n/a | -0.007 | 18.6 |

SP = survival probability; NP = nonparametric. n/a indicates model did not converge or produced unreliable fit.

**Supplementary Figure S1.**
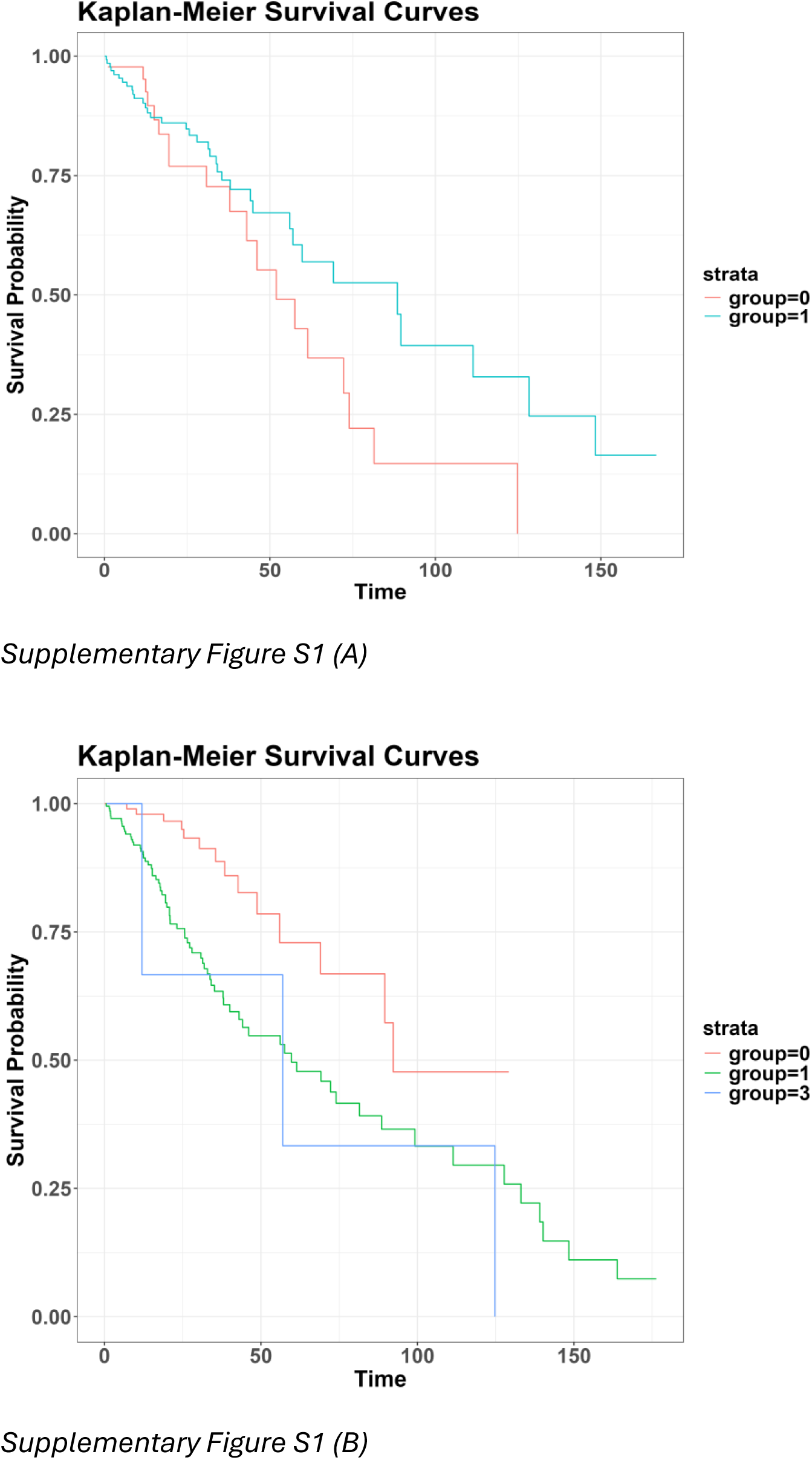
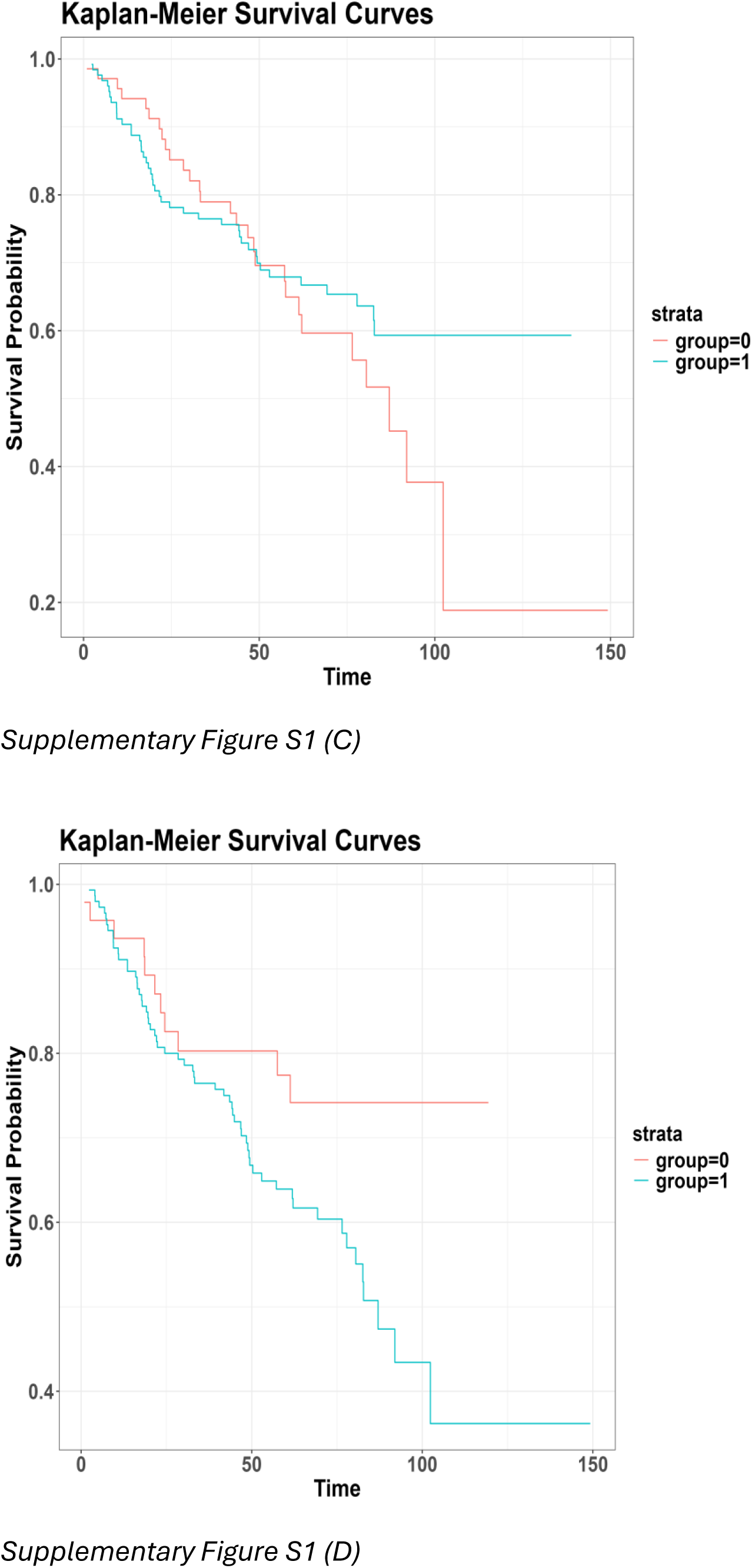
Kaplan-Meier survival curves by treatment group for overall survival. (A) TCGA Chemotherapy. (B) TCGA Radiotherapy. (C) CGGA Chemotherapy. (D) CGGA Radiotherapy. These unadjusted curves illustrate the confounding by indication challenge: in several scenarios, treated patients appear to have worse raw survival due to selection of higher-risk patients for treatment.

**Supplementary Figure S2.**
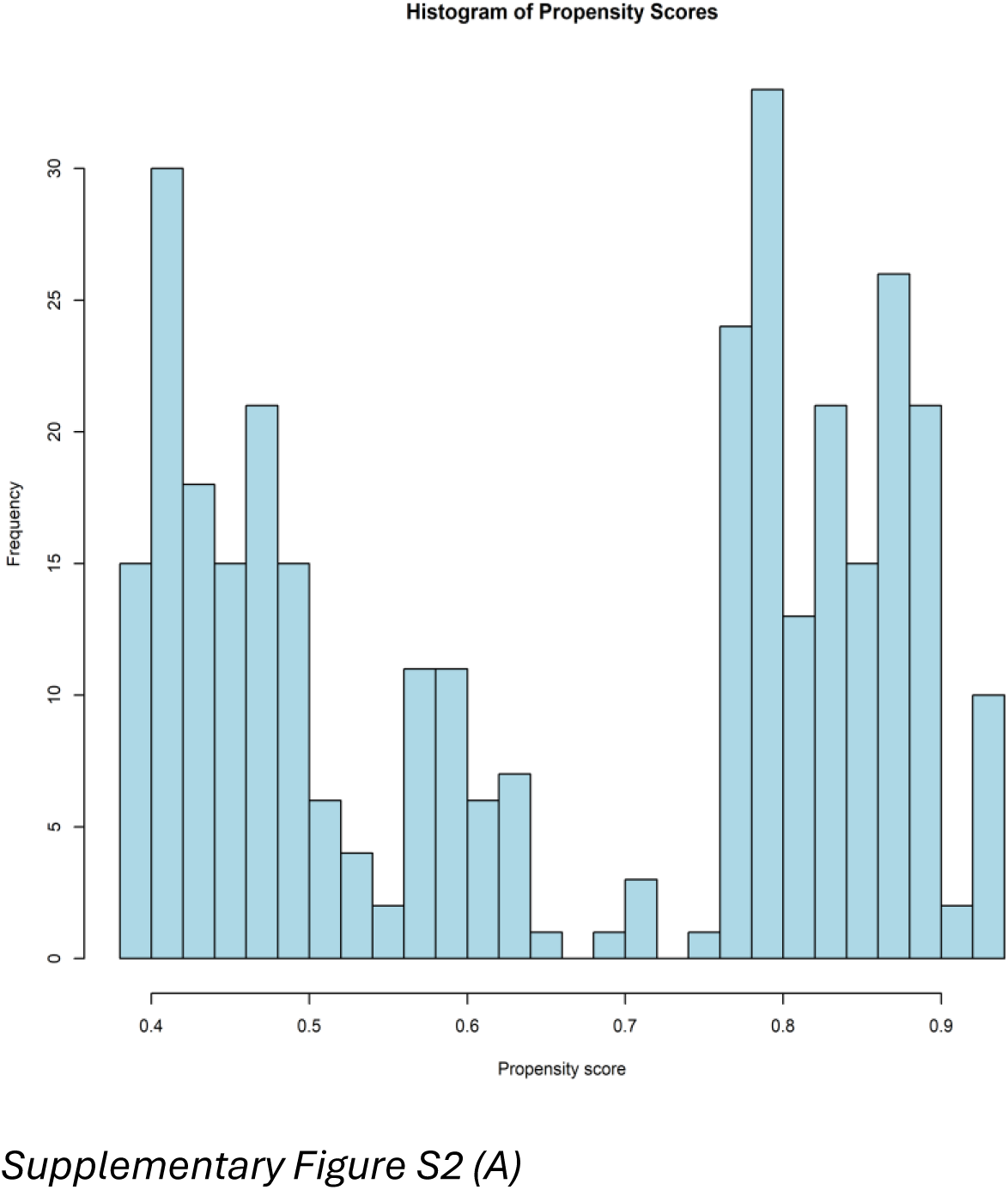
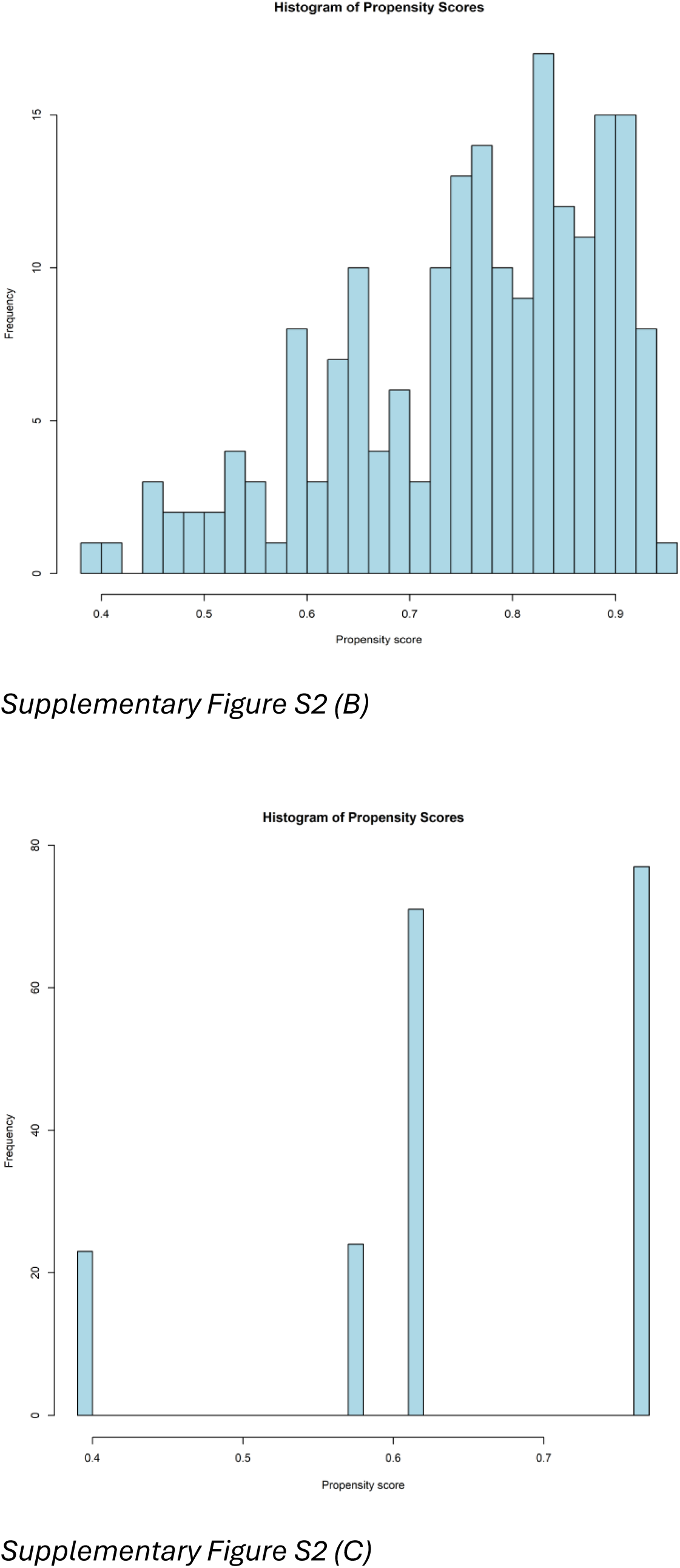
Propensity score distributions by treatment group. (A) TCGA Radiotherapy. (B) CGGA Radiotherapy. (C) CGGA Chemotherapy. Histograms show estimated propensity scores for treated (blue) and untreated (red) patients in the training set.

**Supplementary Figure S3.**
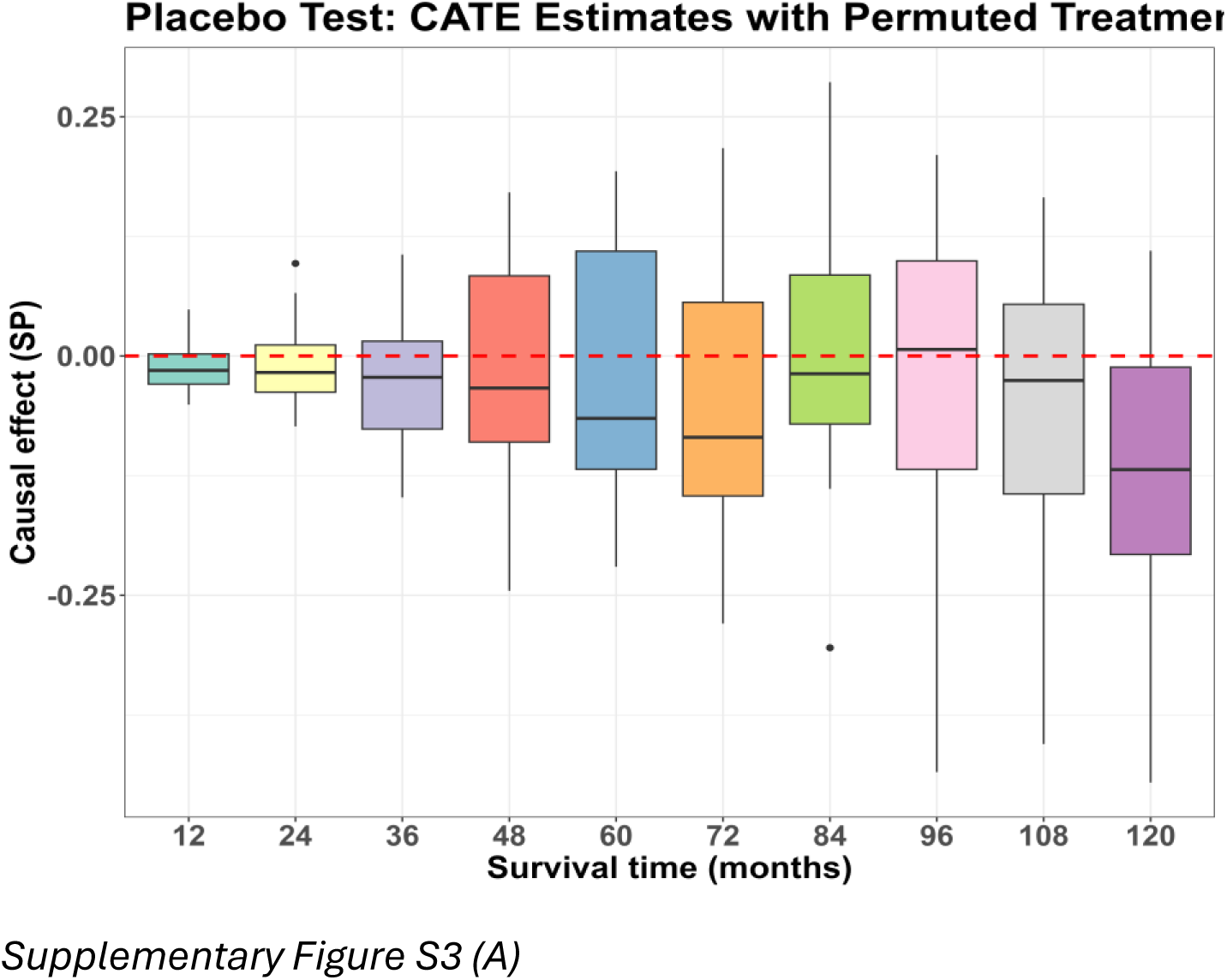
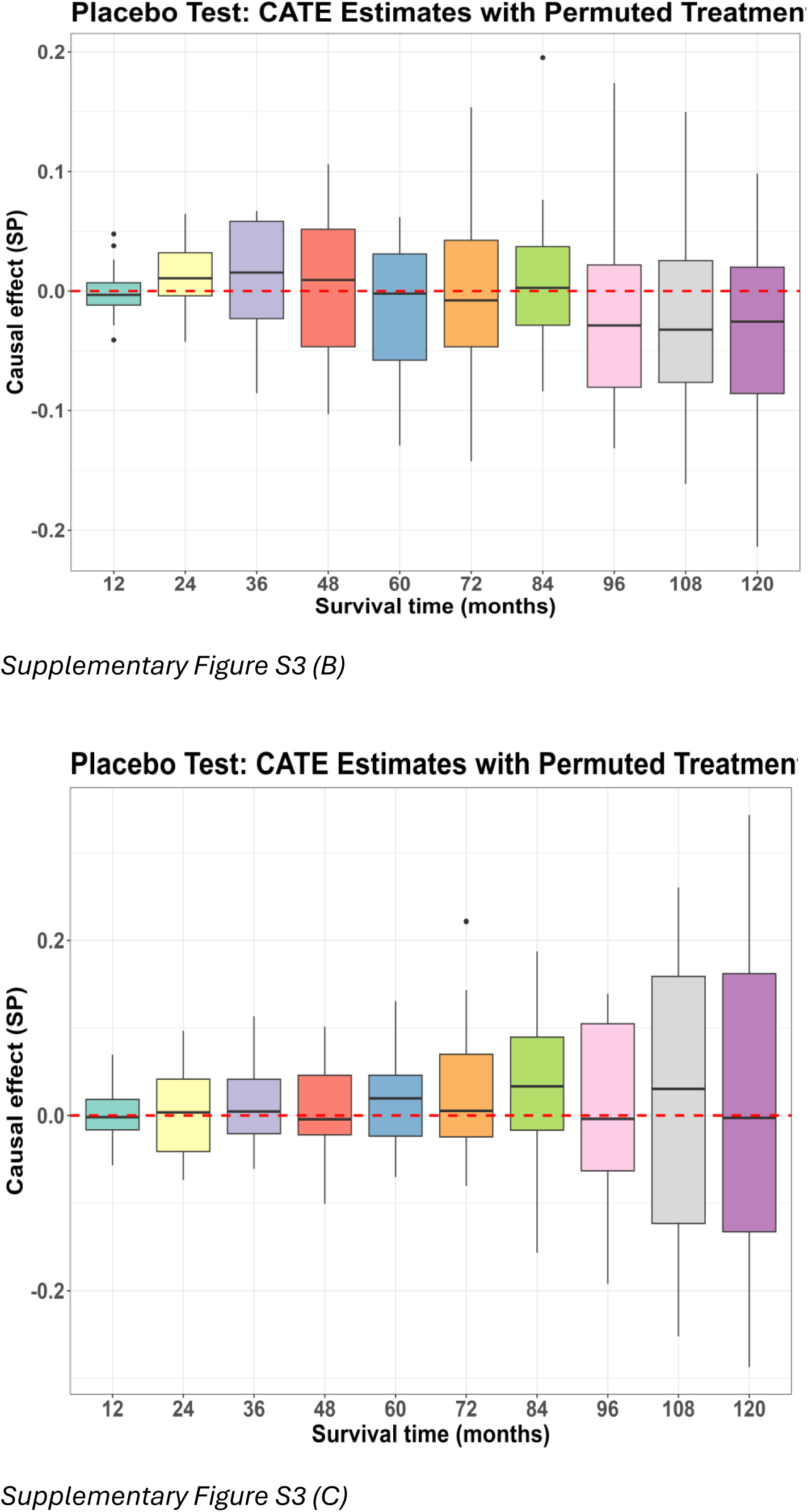
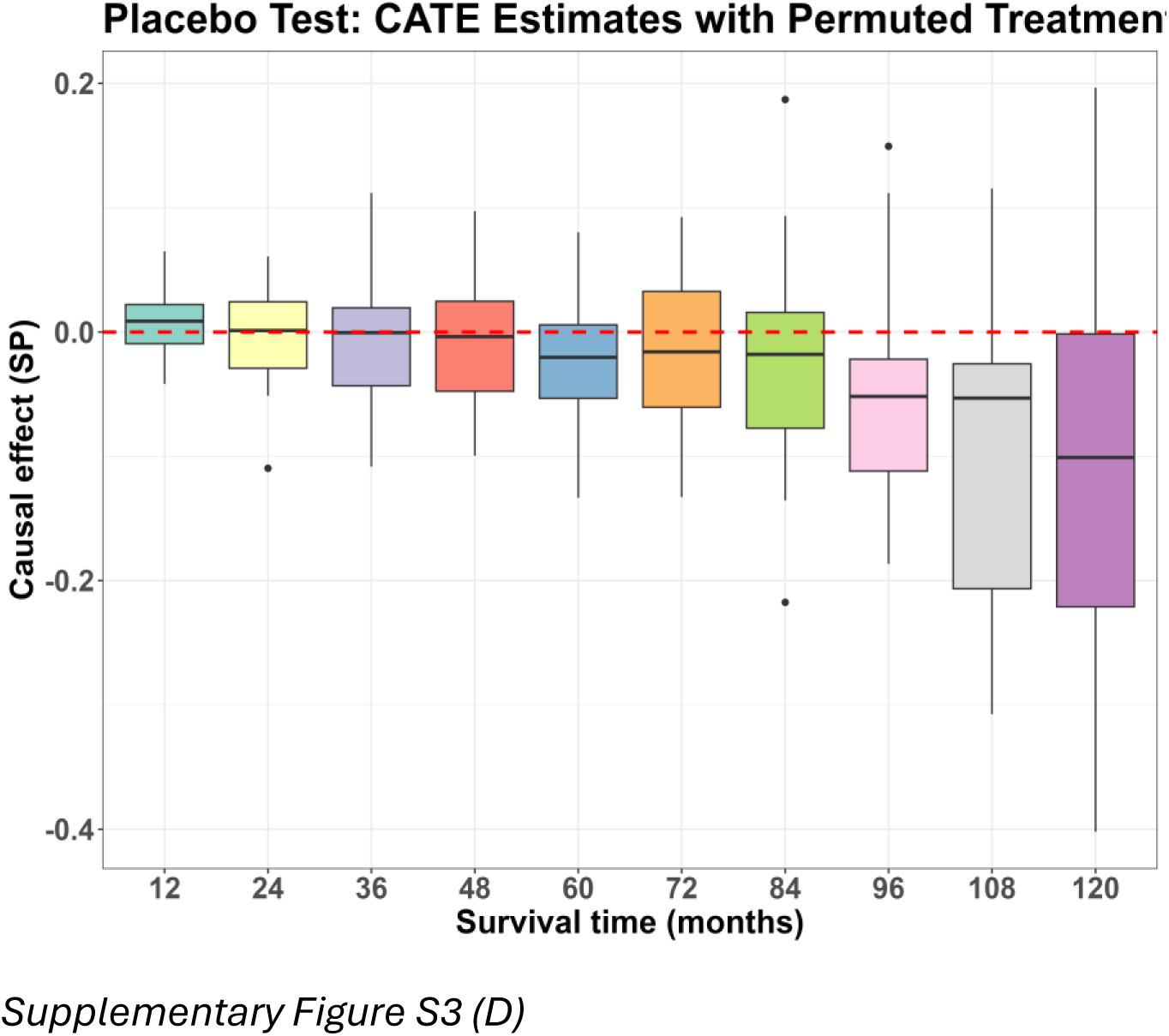
Placebo (treatment permutation) test results for the survival probability metric. (A) TCGA Chemotherapy OS. (B) TCGA Radiotherapy OS. (C) CGGA Chemotherapy OS. (D) CGGA Radiotherapy OS. Boxplots show the distribution of ATE estimates across 20 permutation replicates at each time horizon. Dashed horizontal line at zero indicates the expected null effect.

**Supplementary Figure S4.**
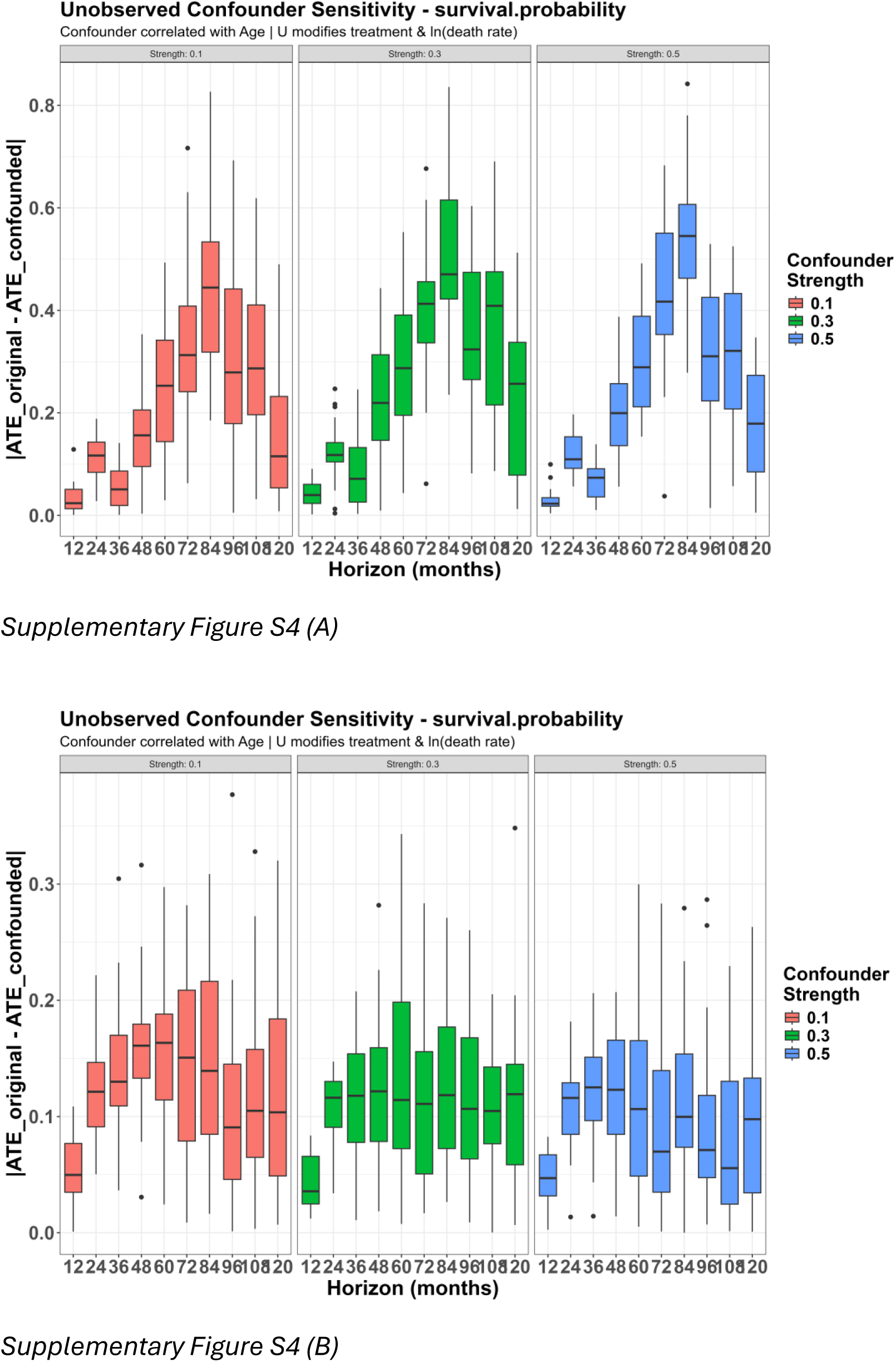
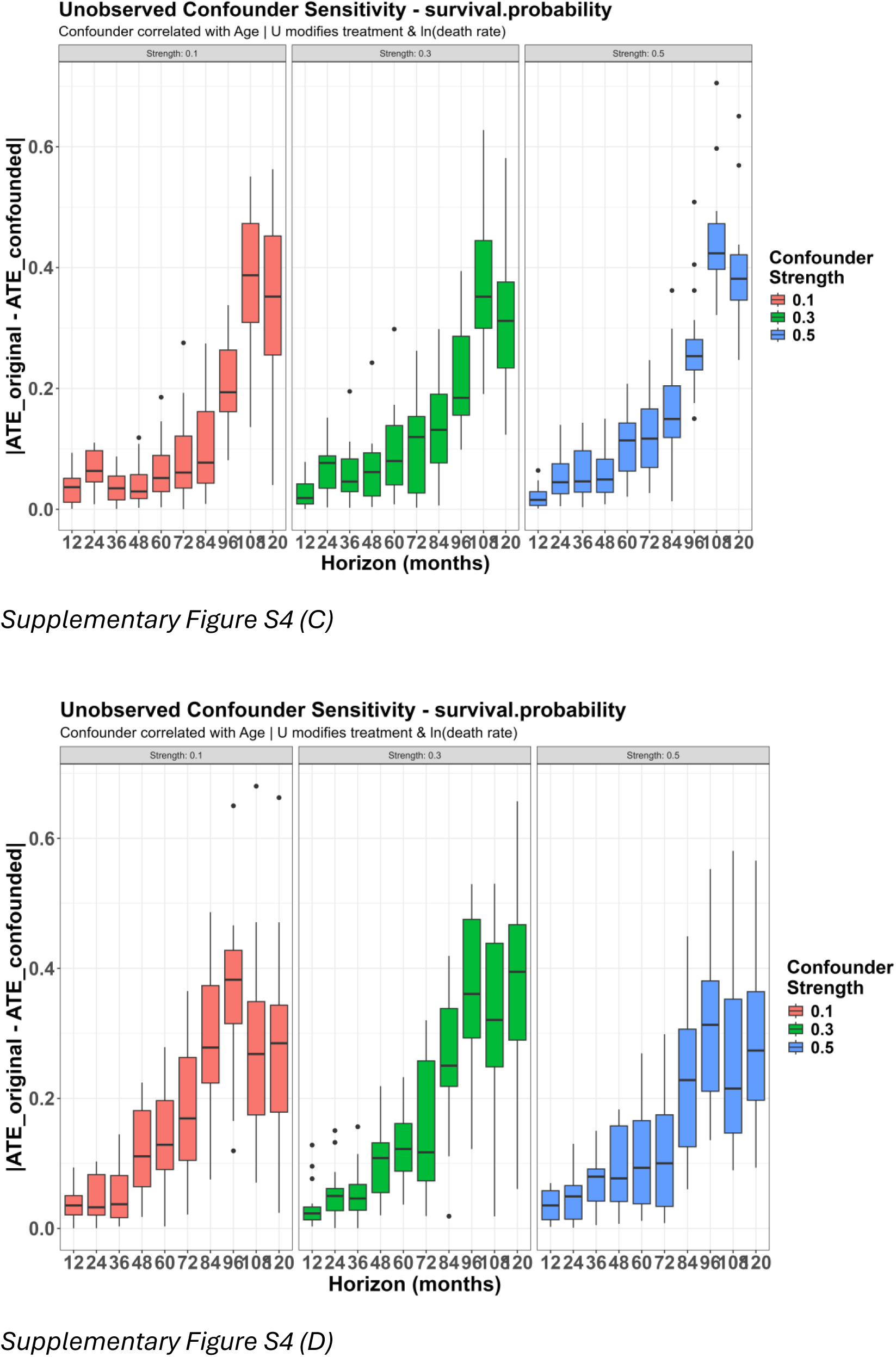
Unobserved confounder sensitivity test results for the survival probability metric. (A) TCGA Chemotherapy OS. (B) TCGA Radiotherapy OS. (C) CGGA Chemotherapy OS. (D) CGGA Radiotherapy OS. Each panel shows ATE estimates at varying confounder strengths (gamma = 0.1, 0.3, 0.5) compared to the original estimate.

**Supplementary Figure S5.**
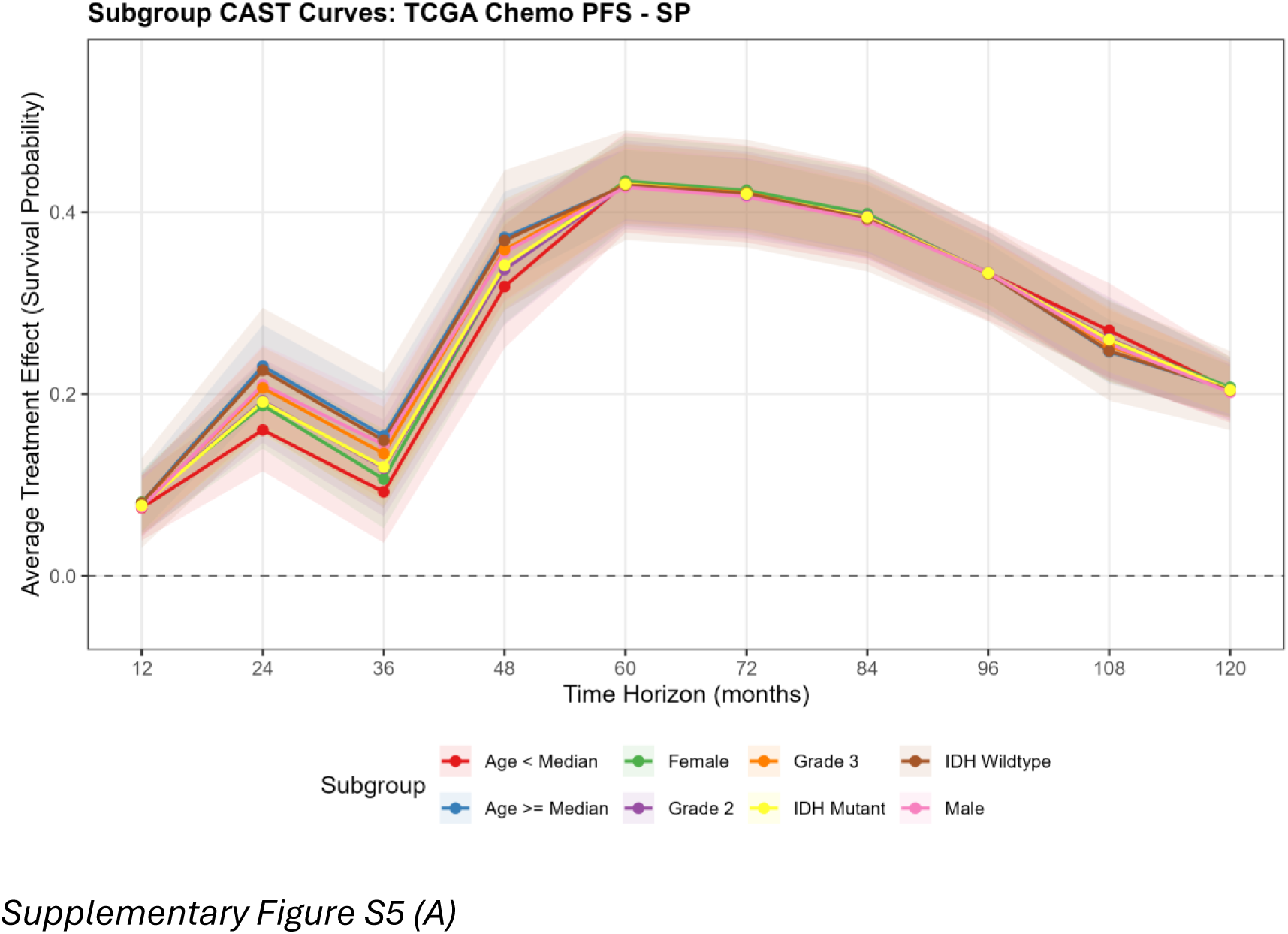
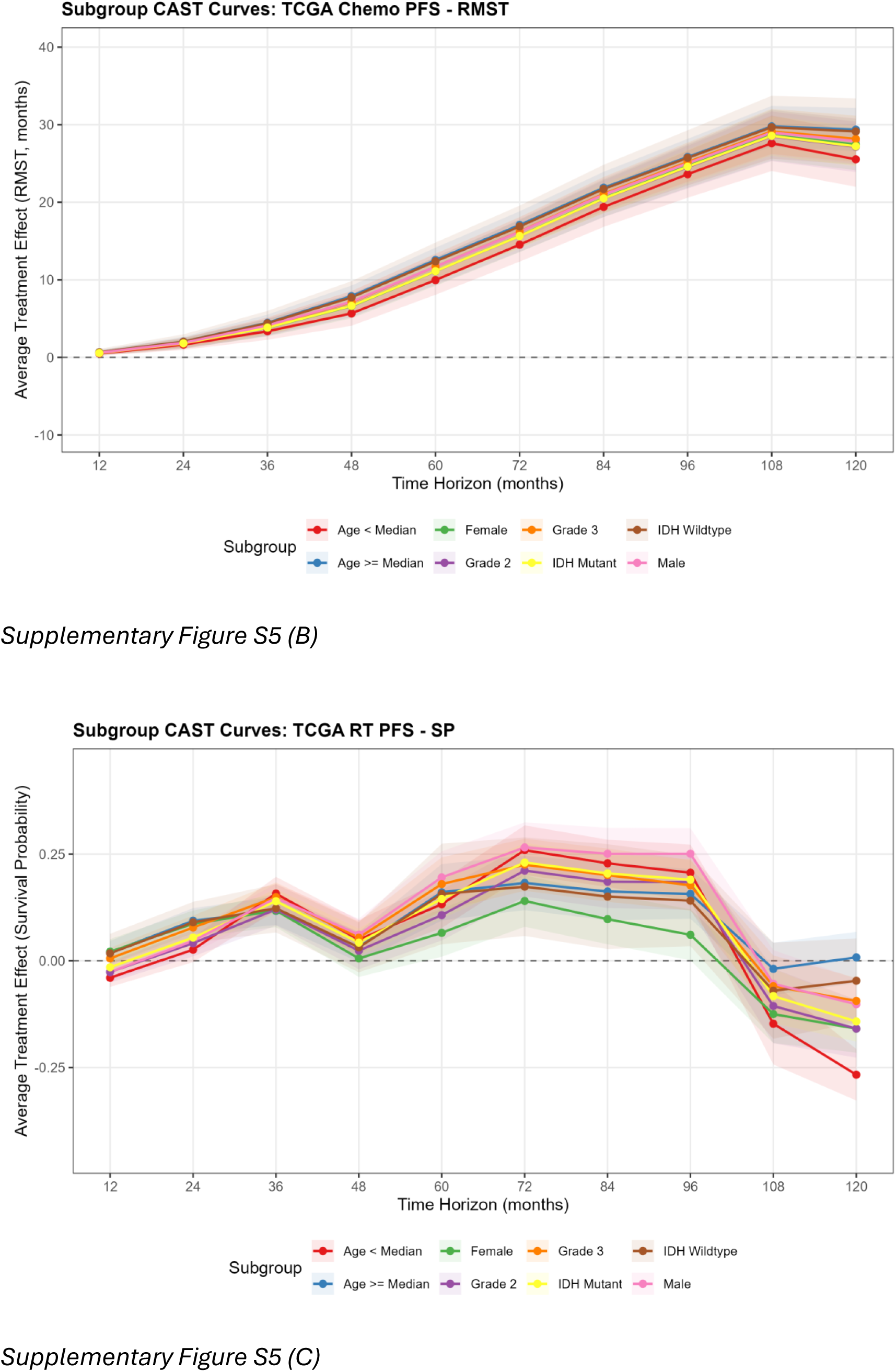
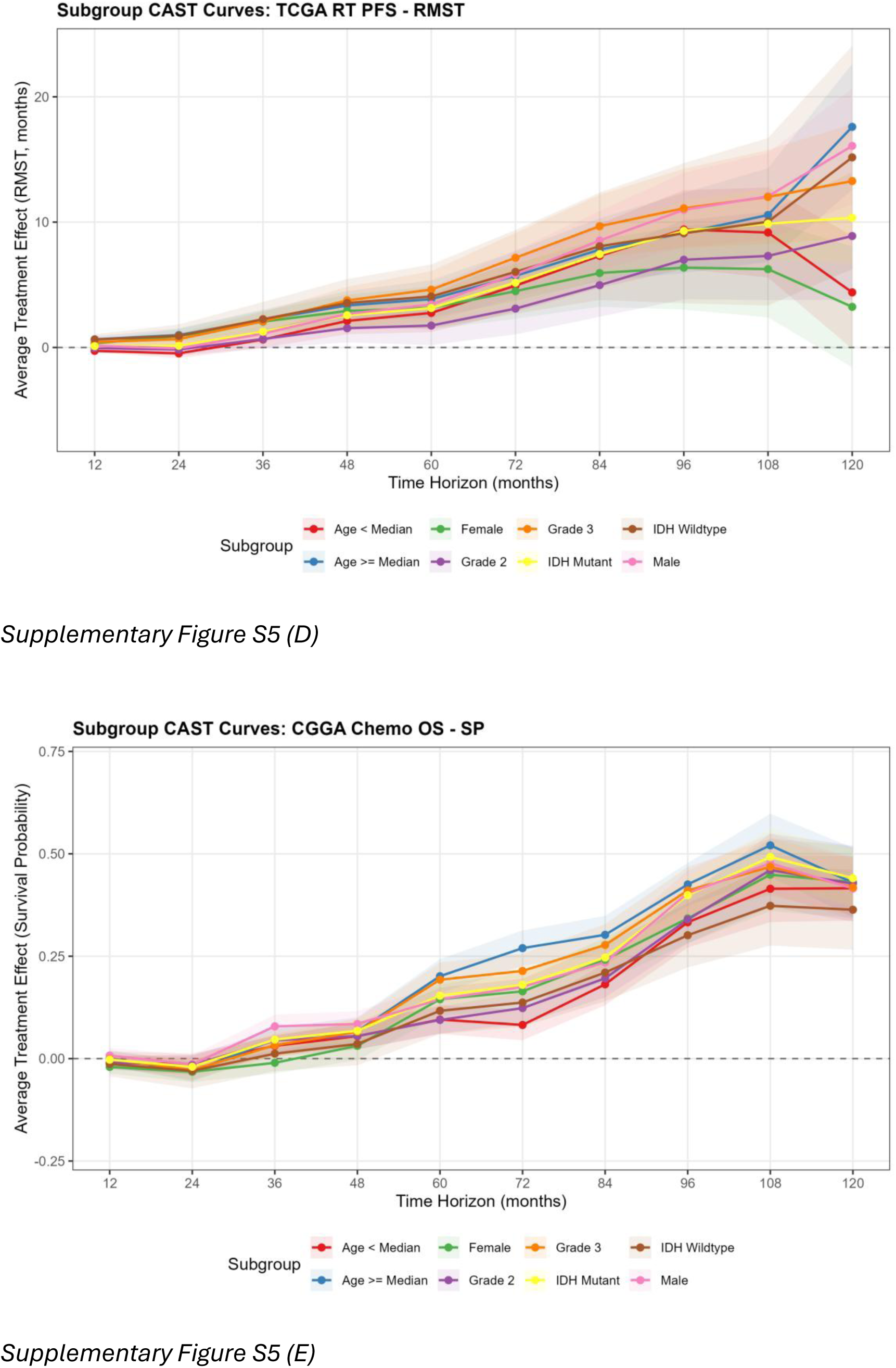
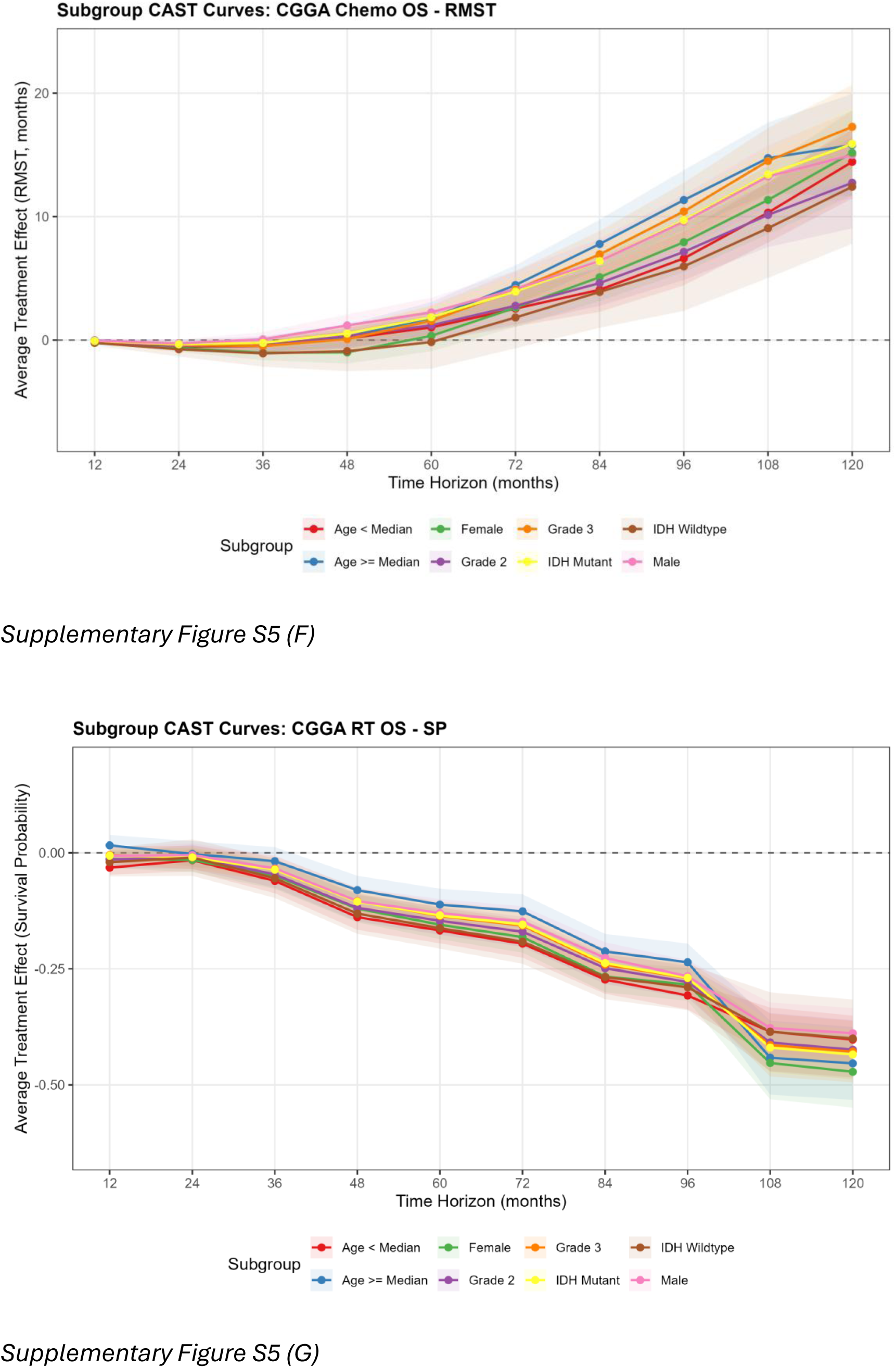
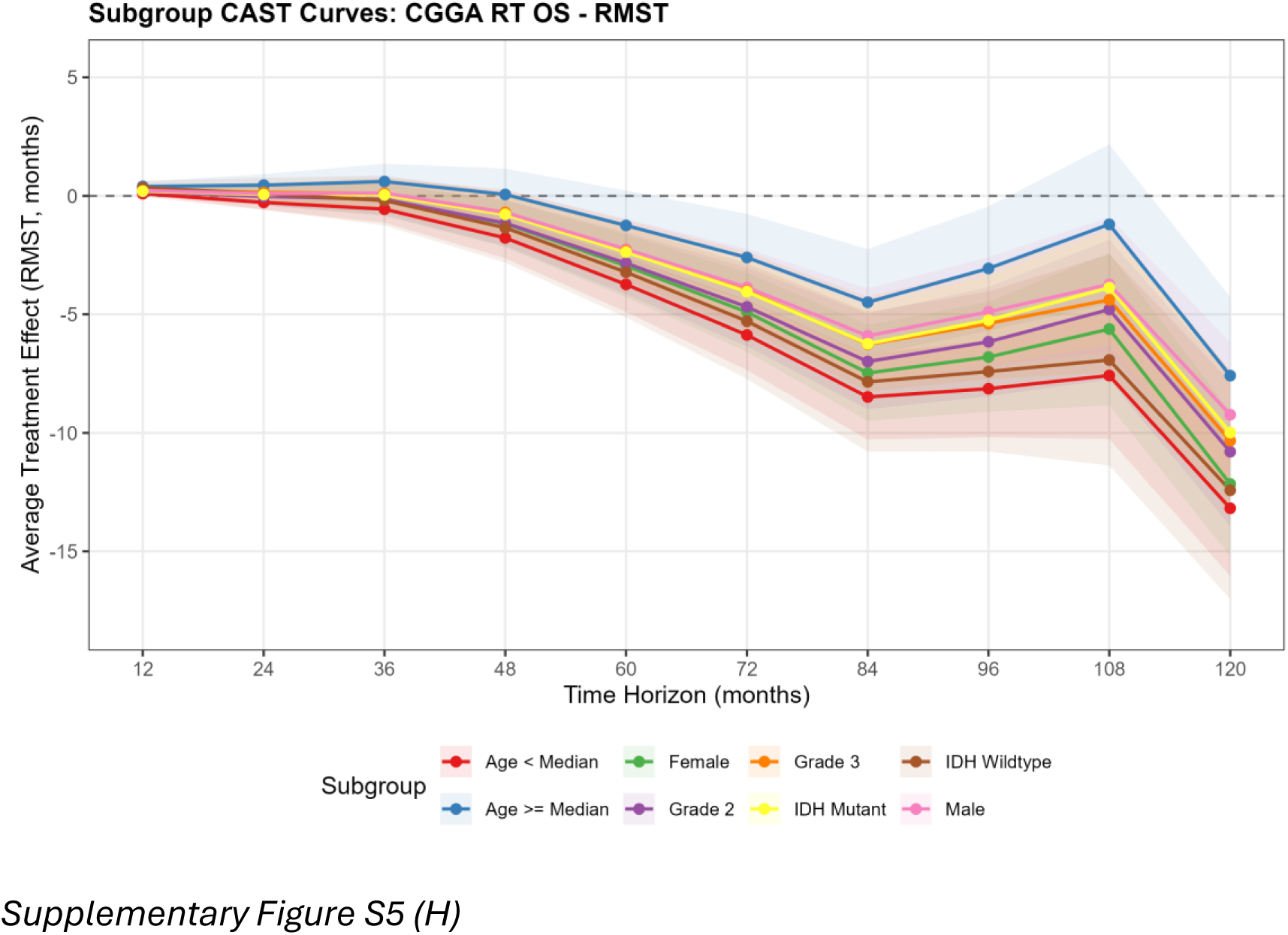
Additional subgroup CAST curves. (A,B) TCGA chemotherapy on PFS: SP and RMST. (C,D) TCGA radiotherapy on PFS: SP and RMST. (E,F) CGGA chemotherapy on OS: SP and RMST. (G,H) CGGA radiotherapy on OS: SP and RMST. Y-axes are zoomed; error bars extending beyond the plot limits are clipped.

## References

1. Bell EH, Zhang P, Shaw EG, et al. Comprehensive Genomic Analysis in NRG Oncology/RTOG 9802: A Phase III Trial of Radiation Versus Radiation Plus Procarbazine, Lomustine (CCNU), and Vincristine in High-Risk Low-Grade Glioma. J Clin Oncol. 2020;38(29):3407–3417.

2. Jonsson P, Lin AL, Young RJ, et al. Genomic Correlates of Disease Progression and Treatment Response in Prospectively Characterized Gliomas. Clin Cancer Res. 2019;25(18):5537–5547.

3. Louis DN, Perry A, Wesseling P, et al. The 2021 WHO Classification of Tumors of the Central Nervous System: a summary. Neuro Oncol. 2021;23(8):1231–1251.

4. Byun YH, Park CK. Classification and Diagnosis of Adult Glioma: A Scoping Review. Brain Neurorehabilitation. 2022;15:e19.

5. Claus EB, Walsh KM, Wiencke JK, et al. Survival and low-grade glioma: the emergence of genetic information. Neurosurg Focus. 2015;38(1):E6.

6. Brat DJ, Verhaak RG, Aldape KD, et al.; Cancer Genome Atlas Research Network. Comprehensive, Integrative Genomic Analysis of Diffuse Lower-Grade Gliomas. N Engl J Med. 2015;372(26):2481–2498.

7. Nakasu S, Deguchi S, Nakasu Y. IDH wild-type lower-grade gliomas with glioblastoma molecular features: a systematic review and meta-analysis. Brain Tumor Pathol. 2023;40(3):143–157.

8. Weller M, van den Bent MJ, Preusser M, et al. EANO guidelines on the diagnosis and treatment of diffuse gliomas of adulthood. Nat Rev Clin Oncol. 2021;18(3):170–186.

9. Lombardi G, Barresi V, Indraccolo S, et al. Clinical Management of Diffuse Low-Grade Gliomas. Cancers. 2020;12(10):3008.

10. Buckner JC, Shaw EG, Pugh SL, et al. Radiation plus Procarbazine, CCNU, and Vincristine in Low-Grade Glioma (RTOG 9802). N Engl J Med. 2016;374(14):1344–1355.

11. Cairncross G, Wang M, Shaw E, et al. Phase III trial of chemoradiotherapy for anaplastic oligodendroglioma: long-term results of RTOG 9402. J Clin Oncol. 2013;31(3):337–343.

12. van den Bent MJ, Brandes AA, Taphoorn MJ, et al. Adjuvant procarbazine, lomustine, and vincristine chemotherapy in newly diagnosed anaplastic oligodendroglioma: long-term follow-up of EORTC brain tumor group study 26951. J Clin Oncol. 2013;31(3):344–350.

13. van den Bent MJ, Tesileanu CMS, Wick W, et al. Adjuvant and concurrent temozolomide for 1p/19q non-co-deleted anaplastic glioma (CATNON; EORTC study 26053-22054): second interim analysis of a randomised, open-label, phase 3 study. Lancet Oncol. 2021;22(6):813–823.

14. Lassman AB, Iwamoto FM, Cloughesy TF, et al. International retrospective study of over 1000 adults with anaplastic oligodendroglial tumors. Neuro Oncol. 2011;13(6):649–659.

15. Kacimi SEO, Saidi-Ahmed S, Houillier C, et al. Survival Outcomes Associated With First-Line Procarbazine, CCNU, and Vincristine or Temozolomide in Combination With Radiotherapy in IDH-Mutant 1p/19q-Codeleted Grade 3 Oligodendroglioma. J Clin Oncol. 2025;43(3):329–338.

16. Jaeckle KA, Ballman KV, van den Bent MJ, et al. CODEL: phase III study of RT, RT + TMZ, or TMZ for newly diagnosed 1p/19q codeleted oligodendroglioma. Analysis from the initial study design. Neuro Oncol. 2021;23(3):457–467.

17. Schiff D, Ahluwalia M, Engelman D, et al. Randomized phase III study of radiotherapy with or without temozolomide for low-grade glioma: ECOG-ACRIN E3F05. Presented at: Society for Neuro-Oncology Annual Meeting; November 2024; Houston, TX.

18. Laack NN, Sarkaria JN, Buckner JC. Radiation Therapy Oncology Group 9802: Controversy or Consensus in the Treatment of Newly Diagnosed Low-Grade Glioma? Semin Radiat Oncol. 2015;25(3):197–202.

19. Rosenbaum PR, Rubin DB. The central role of the propensity score in observational studies for causal effects. Biometrika. 1983;70(1):41–55.

20. Athey S, Tibshirani J, Wager S. Generalized random forests. Ann Stat. 2019;47(2):1148–1178.

21. Cui Y, Kosorok MR, Sverdrup E, Wager S, Zhu R. Estimating heterogeneous treatment effects with right-censored data via causal survival forests. J R Stat Soc Ser B. 2023;85(2):179–211.

22. Hu L, Ji J, Li F. Estimating heterogeneous survival treatment effect in observational data using machine learning. Stat Med. 2021;40(21):4691–4713.

23. Yang E, Vasishtha R, Dad LK, Kachnic LA, Hope A, Wang E, Wu X, Yuan Y, Brenner DJ, Shuryak I. CAST: Time-Varying Treatment Effects with Application to Chemotherapy and Radiotherapy on Head and Neck Squamous Cell Carcinoma. *arXiv*. 2025. arXiv:2505.06367. Presented at: NeurIPS 2025 Workshops (AI4Science, CauScien, TS4H); December 6-7, 2025; San Diego, CA.

24. Ledoit O, Wolf M. A well-conditioned estimator for large-dimensional covariance matrices. J Multivar Anal. 2004;88(2):365–411.

25. Li F, Morgan KL, Zaslavsky AM. Balancing covariates via propensity score weighting. J Am Stat Assoc. 2018;113(521):390–400.

26. VanderWeele TJ, Ding P. Sensitivity Analysis in Observational Research: Introducing the E-Value. Ann Intern Med. 2017;167(4):268–274.

27. Textor J, van der Zander B, Gilthorpe MS, Liskiewicz M, Ellison GT. Robust causal inference using directed acyclic graphs: the R package ‘dagitty’. Int J Epidemiol. 2016;45(6):1887–1894.

28. Mo Z, Zhang Y, Cao Z, et al. Epidemiological characteristics and genetic alterations in adult diffuse glioma in East Asian populations. Cancer Biol Med. 2022;19(10):1440–1459.

29. Poon MT, Woo PY, Helali AE, Brennan PM. Over one year of survival difference between distinct cohorts of glioblastoma patients. medRxiv. 2023. doi:10.1101/2023.03.20.23287451.

30. Tunthanathip T, Madteng S. Effect of the Extent of Resection on Survival Outcome in Glioblastoma: Propensity Score Approach. Arq Bras Neurocir. 2020;40(1):e27–e35.

31. Zhao Z, Zhang KN, Wang Q, et al. Chinese Glioma Genome Atlas (CGGA): A Comprehensive Resource with Functional Genomic Data from Chinese Glioma Patients. Genomics Proteomics Bioinformatics. 2021;19(1):1–12.

32. Fisher BJ, Hu C, Macdonald DR, et al. Phase 2 study of temozolomide-based chemoradiation therapy for high-risk low-grade gliomas: preliminary results of Radiation Therapy Oncology Group 0424. Int J Radiat Oncol Biol Phys. 2015;91(3):497–504.

33. Baumert BG, Hegi ME, van den Bent MJ, et al. Temozolomide chemotherapy versus radiotherapy in high-risk low-grade glioma (EORTC 22033-26033): a randomised, open-label, phase 3 intergroup study. Lancet Oncol. 2016;17(11):1521–1532.

34. Nakamura M, Konishi N, Tsunoda S, et al. Analysis of Prognostic and Survival Factors Related to Treatment of Low-Grade Astrocytomas in Adults. Oncology. 2000;58(2):108–116.

35. Anderson JR, Cain KC, Gelber RD. Analysis of survival by tumor response. J Clin Oncol. 1983;1(11):710–719.

36. Stekhoven DJ, Buhlmann P. MissForest: non-parametric missing value imputation for mixed-type data. Bioinformatics. 2012;28(1):112–118.

37. van Buuren S, Groothuis-Oudshoorn K. mice: Multivariate Imputation by Chained Equations in R. J Stat Softw. 2011;45(3):1–67.

38. Friedman J, Hastie T, Tibshirani R. Regularization paths for generalized linear models via coordinate descent. J Stat Softw. 2010;33(1):1–22.

